# Deep learning aging marker from retinal images unveils sex-specific clinical and genetic signatures

**DOI:** 10.1101/2025.07.29.25332359

**Authors:** Olga Trofimova, Leah Böttger, Sacha Bors, Yating Pan, Bart Liefers, Jose D. Vargas Quiros, Victor A. de Vries, Michael J. Beyeler, David M. Presby, Dennis Bontempi, Janna Hastings, Caroline C. W. Klaver, Sven Bergmann

## Abstract

Retinal fundus images offer a non-invasive window into systemic aging. Here, we fine-tuned a foundation model (RETFound) to predict chronological age from color fundus images in 71,343 participants from the UK Biobank, achieving a mean absolute error of 2.85 years. The resulting retinal age gap, i.e. the difference between predicted and chronological age, was associated with cardiometabolic traits, inflammation, cognitive performance, all-cause mortality, dementia, cancer, and incident cardiovascular disease. Genome-wide analyses identified genes related to longevity, metabolism, neurodegeneration, and age-related eye diseases. Sex-stratified models revealed consistent performance but divergent biological signatures: males had younger - appearing retinas and stronger links to metabolic syndrome, while in females, both model attention and genetic associations pointed to a greater involvement of retinal vasculature. Our study positions retinal aging as a biologically meaningful and sex-sensitive biomarker that can support more personalized approaches to risk assessment and aging-related healthcare.

## Introduction

Aging is strongly linked to disease susceptibility and mortality risk, yet its biological effects manifest at different rates across individuals—some people experience earlier onset of age-related decline, while others remain relatively healthy even at old age. The ability to measure biological age, distinct from chronological age, has therefore become a growing area of interest in biomedical research [1,2]. Estimators of biological age can offer valuable insights into disease susceptibility, predict health outcomes, and help identify individuals who may benefit from early interventions, such as medications or lifestyle changes [1,2].

Previous research has shown that deep learning models can estimate chronological age from retinal color fundus images (CFIs) with high accuracy [3–13]. Attention maps of such models reveal that the predictive signal draws on a range of anatomical features, including blood vessels [4,7,11,12], the macula [4,7,12], and the optic disc [4,11,12], suggesting that age-relevant information is distributed across multiple retinal regions. Importantly, the difference between predicted age and true age, known as retinal age gap (RAG), has emerged as a promising marker of biological aging. Several studies have reported associations between RAG and cardiovascular diseases (CVD) [5,12,14], chronic obstructive pulmonary disease [8], Parkinson’s disease [15], cancer [12], and all-cause mortality [7,12,16], independent of traditional risk factors. Unlike other biological age markers that require more invasive procedures or costly laboratory assays, retinal age can be estimated cheaply and non-invasively from fundus photographs. This makes it a highly accessible and scalable tool for risk stratification in both clinical and population-level health settings.

As interest in biological age estimation grows, so does the need to understand how these estimates may vary across demographic groups, particularly between sexes. Research in neuroscience offers a relevant example: studies using brain imaging to estimate biological age have often found that female brains appear younger than male brains on average [17–19]. However, the findings are not fully consistent, as one study reports no sex differences [20], another finds female brains to appear older [21], and yet another shows region-specific variations in age estimates [22]. Despite such inconsistencies, the overall evidence supports the importance of stratifying analyses by sex, especially given that the brain age gap is differentially associated with disease patterns and risk factors in females and males [19–22]. In contrast, retinal age studies to date have largely relied on models trained on combined male and female samples, leaving it unclear whether, and how, RAG and its clinical correlates differ between sexes.

Traditionally, deep learning in biomedical imaging has relied on convolutional neural networks (CNNs), but recent advances have introduced Vision Transformers as viable alternatives. While there is no consensus on which architecture performs best overall, evidence suggests performance is task- and data-dependent. For instance, Vision Transformers often outperform CNNs in classification tasks when large datasets and pretraining are available, whereas CNN-based models like nnU-Net continue to excel in 3D medical image segmentation, particularly when rigorously validated and carefully configured [23,24]. RETFound, a foundation model for retinal images pretrained using a masked autoencoder with a large Vision Transformer backbone, has shown strong performance when fine-tuned for disease classification tasks, outperforming other contrastive learning-based approaches [25]. However, RETFound has not yet been applied to age prediction or explored in the context of retinal aging.

In this study, we aimed to extend and deepen the current understanding of retinal aging through several complementary analyses (**Fig. 1**). First, we benchmarked the performance of RETFound in age prediction from CFIs against conventional CNNs. Second, we trained sex-specific models—using separate cohorts of females and males—to investigate whether modeling each sex independently enhanced performance or revealed differences in aging patterns. We then explored the sex-specific value of predicted retinal age and RAG, particularly with respect to disease associations and genetic correlates. Third, we sought to interpret the internal representations of the model by analyzing attention maps and comparing the model’s latent space to explicit image features. Through these analyses, we aim to uncover the retinal correlates of biological aging, assess their clinical utility, and understand how they may differ between females and males.

**Figure 1.**
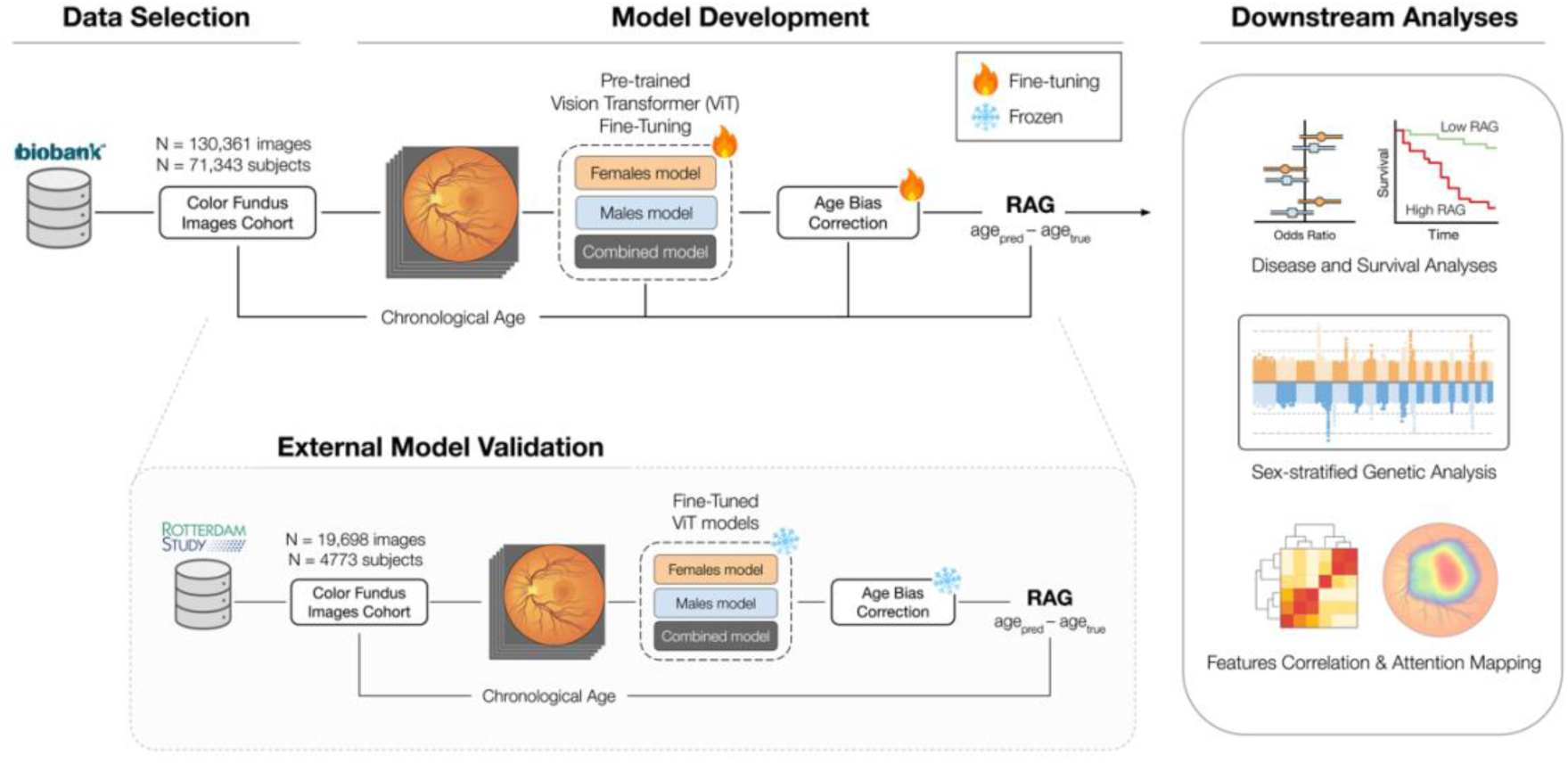
Study overview. We selected a subset of UK Biobank participants with retinal color fundus images that passed quality control. Using 5-fold cross-validation, we fine-tuned RETFound to predict age in a combined sample as well as in male- and female-specific samples. After correcting for regression-to-the-mean bias, we computed the retinal age gap (RAG). We conducted downstream analyses including disease associations, survival analysis, genome-wide association study, correlations between latent and explicit features, and model attention mapping, in both the combined and sex-specific samples. We then tested model performance and replication of genetic associations in an external dataset from the Rotterdam Study.

## Results

### RETFound predicts age with high accuracy and generalizes to an external dataset

Fine-tuning RETFound for age prediction yielded consistent performance across the five models trained via five-fold cross-validation (see **Table S1** for details). After pooling out-of-sample predictions across all folds (*n* = 130,360 fundus images from the UK Biobank (UKB)), the mean absolute error (MAE) was 2.99 years, Pearson’s correlation (*r*) between predictions and chronological ages was 0.88, and the coefficient of determination (R²) was 0.78 (**Table 1**). When averaging predictions across all available images from each participant at a given time point, the MAE improved to 2.85 years, with an *r* of 0.90 and an R² of 0.80. The model trained on a female-specific cohort achieved an MAE of 3.05 years, an *r* of 0.88, and an R² of 0.78 before averaging across images, which improved to an MAE of 2.92, an *r* of 0.89, and an R² of 0.80 after averaging. Similarly, the male-specific model showed an MAE of 3.09, an *r* of 0.88, and an R² of 0.77 before averaging and improved to an MAE of 2.96, an *r* of 0.89, and an R² of 0.79 after averaging. The MAE difference between female and male models was not significant (*z* = -0.007, *p* = 0.99 for both image- and subject-level MAE).

**Table 1.** Model performance in the UK Biobank out-of-sample test sets and in the Rotterdam Study (external validation). Performance for female retinas evaluated on male-trained models, and vice versa, is also shown after applying age bias correction, to account for potential age distribution differences between females and males. MAE: mean absolute error, r: Pearson’s correlation coefficient, R^2^: coefficient of determination, RAG: retinal age gap, SD: standard deviation.

| Sex | Predictions | UK Biobank |  |  |  |  | Rotterdam Study |  |  |  |  |
| --- | --- | --- | --- | --- | --- | --- | --- | --- | --- | --- | --- |
| | | N | r | MAE | $R^2$ | RAG mean (SD) | N | r | MAE | $R^2$ | RAG mean (SD) |
| Combined | Image-level | 130,360 | 0.88 | 2.99 | 0.78 | 0.19 (3.86) | 10,354 | 0.83 | 3.58 | 0.61 | -2.07 (4.02) |
|  | Subject-level | 71,343 | 0.90 | 2.85 | 0.80 | 0.18 (3.66) | 4757 | 0.85 | 3.35 | 0.66 | -1.94 (3.73) |
| Female | Image-level | 58,646 | 0.88 | 3.05 | 0.78 | 0.01 (3.94) | 5849 | 0.83 | 3.30 | 0.66 | -1.03 (4.08) |
|  | Subject-level | 32,918 | 0.89 | 2.92 | 0.80 | 0.002 (3.76) | 2688 | 0.85 | 3.07 | 0.70 | -0.88 (3.80) |
|  | Male-trained model, subject-level | 32,918 | 0.88 | 3.07 | 0.77 | 0.64 (3.89) | 2688 | 0.84 | 3.47 | 0.62 | -2.01 (3.88) |
|  | Male-trained model, subject-level, corrected | 32,918 | 0.88 | 3.36 | 0.73 | 0.73<br>(4.27) | 2688 | 0.84 | 3.50 | 0.61 | 0.67<br>(4.38) |
| Male | Image-level | 58,512 | 0.88 | 3.09 | 0.77 | 0.09<br>(4.00) | 4505 | 0.83 | 3.86 | 0.56 | -2.68<br>(4.08) |
|  | Subject-level | 32,918 | 0.89 | 2.96 | 0.79 | 0.07<br>(3.81) | 2069 | 0.85 | 3.61 | 0.61 | -2.51<br>(3.76) |
|  | Female-trained model, subject-level | 32,918 | 0.88 | 3.08 | 0.77 | -0.66<br>(3.89) | 2069 | 0.86 | 3.19 | 0.70 | -1.51<br>(3.69) |
|  | Female-trained model, subject-level, corrected | 32,918 | 0.88 | 3.54 | 0.71 | -0.83<br>(4.41) | 2069 | 0.86 | 3.53 | 0.62 | -0.81<br>(4.36) |

To assess the generalizability of our models beyond the UKB, we tested their performance on independent sets of CFIs from the Rotterdam Study (RS). The image-level MAE was 3.58 years in the combined sample, 3.30 in females, and 3.86 in males (see **Table 1**). To quantify variability across models, we calculated the mean per-image standard deviation (SD) of the five model predictions, which was 0.99 years in the combined sample (28% of the MAE), 1.19 years in females (36% of the MAE), and 1.12 years in males (29% of the MAE). The subject-level MAE was 3.35 years in the combined sample, 3.07 years in females, and 3.61 years in males. The MAE was not significantly different between sexes, neither at the image level (*z* = -0.097, *p* = 0.92), nor at the subject level (*z* = -0.101, *p* = 0.92). While the squared correlation between true and predicted ages was 0.72 for the three models, the R² values were notably lower (0.61 – 0.70), reflecting a systematic bias in age prediction. The mean RAG values were -2.51 years in males, -0.88 years in females, and -1.94 years in the combined sample, indicating that the model consistently underestimated age in the RS, especially in males.

In both the UKB and the RS, predicted age tended to be overestimated in younger individuals and underestimated in older individuals (**Fig. S1a**), a regression-to-the-mean pattern commonly observed in aging biomarker studies [26]. Therefore, we applied the bias correction method proposed by Cole and colleagues [18], which rendered predicted age error orthogonal to chronological age (see **Fig. S1b**).

### Retinal age gap is consistently lower in males

In the UKB combined cohort, males had a lower RAG than females on average (Δ = 0.28, *t* = 8.9, *p* = 7e-19, Cohen’s *d* = 0.069; see **Fig. S2a**), indicating that the model estimated male retinas as appearing younger, though the effect was small. Additionally, cross-application of sex-specific models revealed that male retinas evaluated using the female-trained model had a mean RAG of -0.83, whereas female retinas evaluated using the male-trained model had a mean RAG of 0.73 after applying age bias correction (see **Table 1**), consistent with the observation that the models tended to estimate male retinas as appearing younger than female retinas. Given this somewhat unexpected sex difference, opposite to what might be anticipated based on lifespan trends, we explored whether it could be explained by anatomical differences, such as overall ocular size. Since body height is known to correlate with ocular dimensions [27], we regressed standing height out of the RAG. This adjustment attenuated the sex difference in RAG, which nonetheless remained significant (Δ = 0.16, *t* = 5.0, *p* = 6e-07, Cohen’s *d* = 0.040; see **Fig. S2b**). Height and height-squared were therefore included as covariates in all subsequent analyses to account for potential confounding.

In the RS, we observed a similar pattern: males had lower mean RAG than females, indicating younger-appearing retinas (Δ = 0.37, *t* = 2.9, *p* = 0.004, Cohen’s *d* = 0.085). When height was regressed out of the RAG, the sex difference was again reduced but not removed (Δ = 0.30, *t* = 2.4, *p* = 0.015, Cohen’s *d* = 0.071). Cross-application of sex-specific models yielded consistent results, with male retinas evaluated using the female-trained model showing a mean RAG of -0.81, and female retinas evaluated using the male-trained model showing a mean RAG of 0.67. These findings replicate the UKB results and further support the observation that retinal age predictions tended to estimate male retinas as appearing younger than female retinas.

### Retinal age gap is associated with metabolic, inflammatory, and ocular health indicators

After adjusting for socio-demographic variables, higher RAG was significantly associated with increased pack-years of smoking, glycated hemoglobin (HbA1c), waist-to-hip ratio, body mass index (BMI), leukocyte count, C-reactive protein (CRP), glucose, triglycerides, systolic blood pressure (SBP), and slower reaction time in the combined sample (**Fig. 2a** and **Table S2** for detailed results). Conversely, RAG was negatively associated with VO₂max, insulin-like growth factor 1 (IGF-1), telomere length, low-density lipoprotein (LDL), high-density lipoprotein (HDL), and total cholesterol. Sex-specific associations generally mirrored the combined results, although males exhibited larger effect sizes across most biomarkers except CRP. Associations found in females but not males comprised neutrophil-to-lymphocyte ratio, LDL, total cholesterol, telomere length, and age at menopause, while male-specific associations included testosterone, SBP, triglycerides, and HDL. Absolute standardized effect sizes ranged from 0.01 to 0.09, reflecting the change in SD units of biomarker levels per SD increase in RAG.

**Figure 2.**
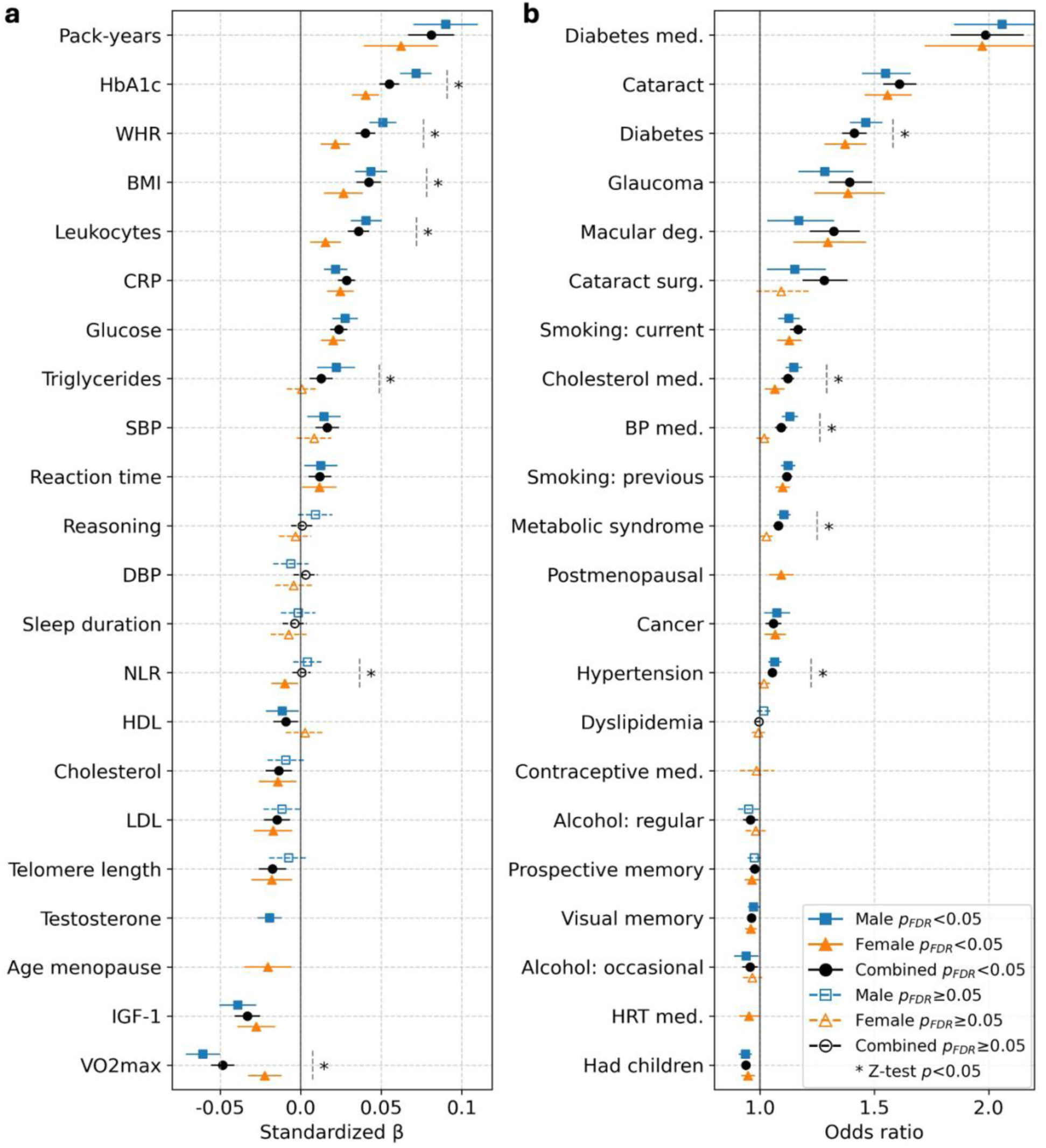
Standardized beta coefficients and odds ratios from models linking retinal age gap to risk factors and diseases. **(a)** Ordinary least square linear regression and **(b)** logistic regression models were adjusted for socio-demographic factors. Outcomes are ordered by average effect size between the three sex categories. Error bars represent the 95% confidence interval. Female- and male-specific effects were compared using a z-test. Sample sizes and detailed results are shown in **Tables S2** and **S3**. BMI: body mass index, BP: blood pressure, CRP: C-reactive protein, DBP: diastolic blood pressure, deg.: degeneration, FDR: false discovery rate, HDL: high-density lipoprotein, HRT: hormone replacement therapy, IGF-1: insulin-like growth factor 1, LDL: low-density lipoprotein, med.: medication, NLR: neutrophil-to-lymphocyte ratio, SBP: systolic blood pressure, surg.: surgery, WHR: waist-to-hip ratio.

For binary health outcomes, in the combined sample, higher RAG was associated with diabetes medication use, a history of cataract, diabetes, glaucoma, macular degeneration, cataract surgery, current or previous smoking status, use of cholesterol or blood pressure medications, metabolic syndrome, cancer (ever diagnosed), and hypertension (**Fig. 2b** and **Table S3**). Negative associations were observed with having children, visual and prospective memory performance, and alcohol consumption. Among females, postmenopausal status was associated with higher RAG, whereas hormone replacement therapy correlated with lower RAG. Males showed stronger associations than females with metabolic traits, including a male-specific association with metabolic syndrome, hypertension, and blood pressure medication. Odds ratios ranged from 0.94 to 2.06 per SD increase in RAG.

### Retinal age gap predicts mortality and incident disease with sex-specific patterns

In survival analyses adjusted for socio-demographics, common cardiovascular risk factors, and previous events of the same category, RAG was significantly associated with all-cause mortality as well as incidence of CVD, chronic obstructive pulmonary disease, stroke, dementia, thrombosis, ischemic heart disease, and cancer in the combined sample (see **Fig. 3** and **Table S4**). When stratified by sex, thrombosis was associated with RAG in females but not males, and hazard ratios (HRs) for CVD and stroke were larger in females, although not significantly. In contrast, RAG predicted dementia and cancer incidence uniquely in males and was more strongly associated with all-cause mortality in males, though confidence intervals were large and the sex difference was only significant for cancer. HRs ranged from 1.03 to 1.20 per SD increase in RAG.

**Figure 3.**
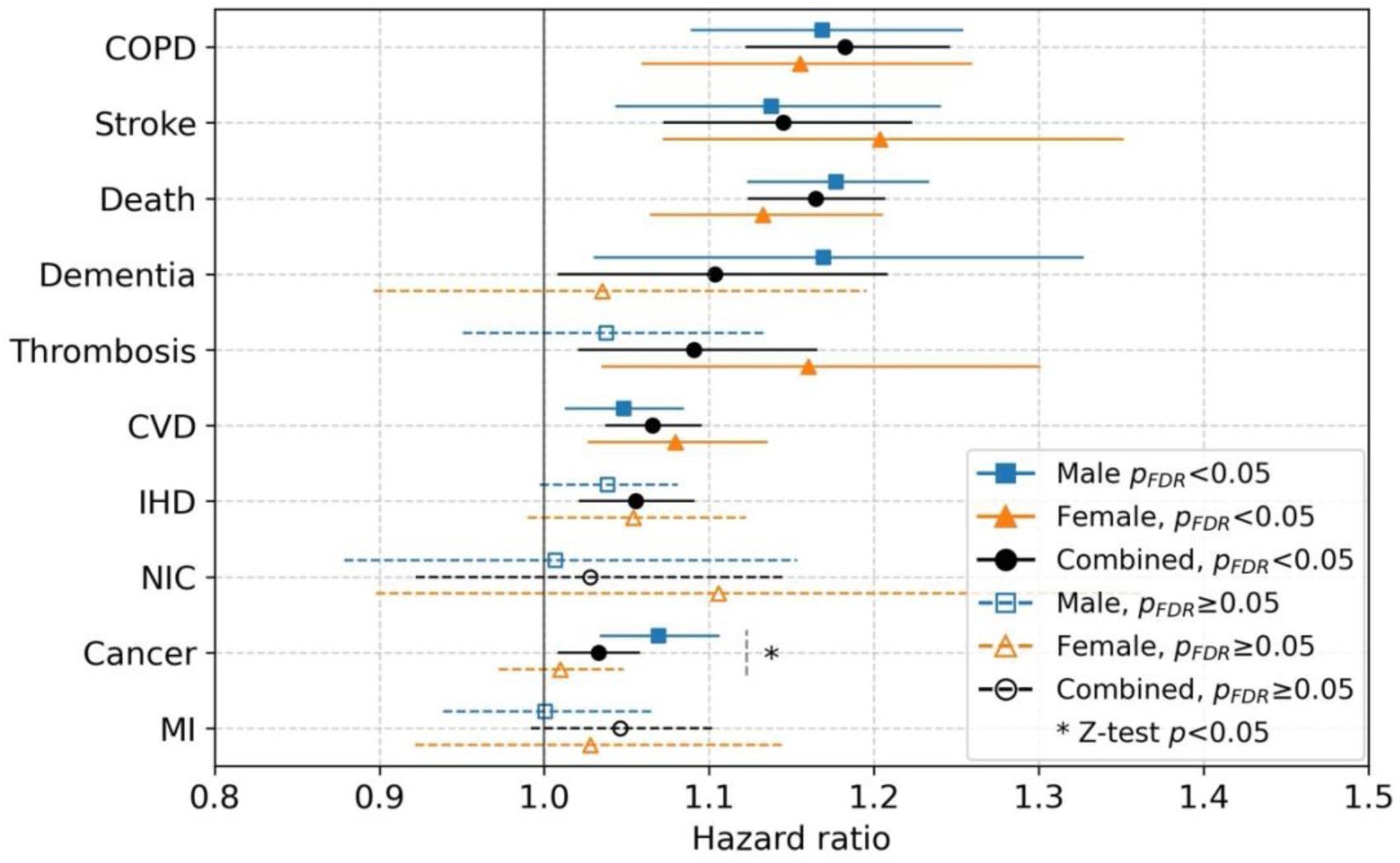
Hazard ratios (HR) from Cox proportional hazards regression analyses linking retinal age gap to mortality and diseases. Models were adjusted for socio-demographic factors, common cardiovascular risk factors, and previous events from the same category. Outcomes are ordered by average HR between the three sex categories. Error bars represent the 95% confidence interval. Female- and male-specific effects were compared using a z-test. Sample sizes, number of events, and detailed results are shown in **Table S4**. COPD: chronic obstructive pulmonary disease, CVD: cardiovascular disease, FDR: false discovery rate, IHD: ischemic heart disease, MI: myocardial infarction, NIC: non-ischemic cardiomyopathy.

### Sex-stratified GWAS reveals shared and distinct genetic associations for retinal age gap

The genome-wide association study (GWAS) of RAG in the UKB identified 40 genome-wide significant loci in the combined sample (**Fig. 4a** and **Table S5**), 10 loci in females (**Fig. 4b**, top and **Table S6**), and 14 loci in males (**Fig. 4b**, bottom and **Table S7**). Gene-wise analysis using PascalX [28] revealed 75 genes across 23 independent linkage disequilibrium (LD) blocks in the combined sample, 18 genes in 5 LD blocks in females, and 16 genes in 5 LD blocks in males, with an overlap of 5 genes between females and males (see **Tables S8–S10**).

**Figure 4.**
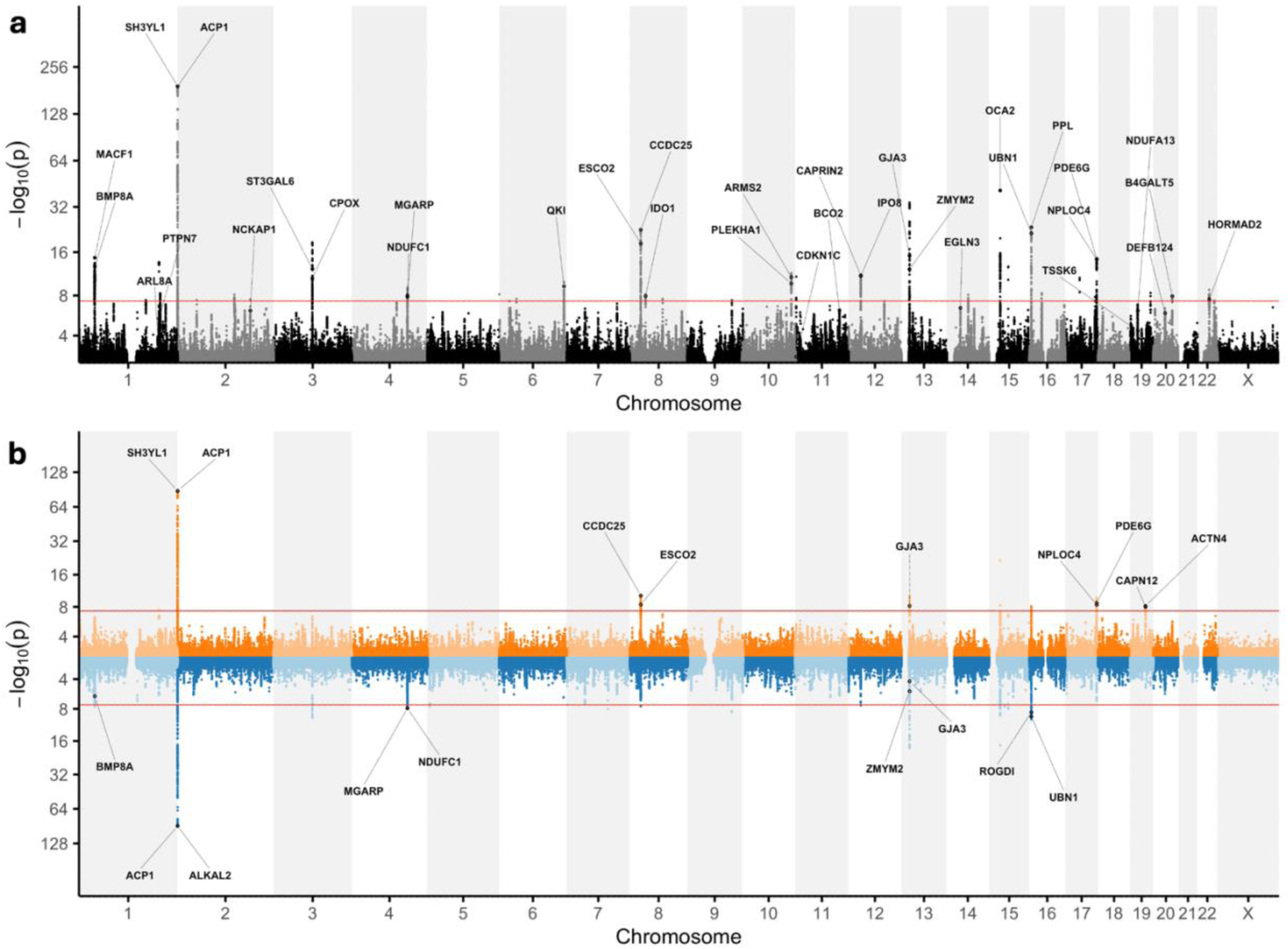
Manhattan plots from the genome-wide association study (GWAS) of retinal age gap, annotated with top genes. **(a)** Manhattan plot of the combined-sample GWAS (n=68,137). **(b)** Miami plot showing the female-specific GWAS (top, orange; n=30,698) and male-specific GWAS (bottom, blue; n=30,958). -log_10_(*p*) values are plotted on a log_2_ scale. The red lines indicate genome-wide significance (5×10^−8^). The annotated genes correspond to those identified as significant by the PascalX gene-based analysis. When more than two genes were identified at the same cytogenetic location, only the top two signals are labeled for clarity (the full gene lists are shown in **Tables S8–S10**).

In the combined sample, the strongest SNP association was observed for rs2290911, mapping within *SH3YL1* on chromosome (chr) 2. Gene-level analysis further revealed prominent signals for *ACP1*, *ALKAL2*/*FAM150B*, and *FAM110C*, which are located in close proximity to *SH3YL1* within the 2p25.3 region. The next strongest signal was found for rs1800407 on chr 15, located within *OCA2*. This was followed by rs11840593 on chr 13, in proximity to *GJA3*. Other notable signals included rs75804753 on chr 16 near *UBN1* and *PPL*, rs13267051 on chr 8 near *CCDC25* and *ESCO2*, rs10935473 on chr 3 within *ST3GAL6*, rs61779328 on chr 1 near *BMP8A* and *MACF1*, and rs3432 near *PDE6G* and *NPLOC4*. The sex-specific analyses revealed distinct genetic associations not observed in the combined sample. In females, unique signals included variants rs12714397 (chr2) and rs8182571 (chr19), and genes *CAPN12* and *ACTN4* (both on chr 19), the latter featuring the variant rs8182571. In contrast, rs79665333 (chr5), rs145132834 (chr7), and rs28844699 (chr15) were identified exclusively in males. Complete results are reported in **Tables S2–7**, and quality control plots for the GWAS analyses are presented in **Fig. S3**. The genomic control factor (λGC) was 1.14, 1.06, and 1.07 for the combined, female, and male GWAS, respectively, indicating modest inflation in the combined analysis and well-calibrated test statistics in the sex-stratified analyses.

The SNP-based heritability (h^2^ ) of RAG, estimated using GREML, was 0.277 (± 0.009) for the combined sample, 0.278 (± 0.019) for females, and 0.247 (± 0.018) for males. The sex difference in h^2^ was not significant (z = 1.15, p = 0.25).

In the RS, 7 of 29 SNPs (11 unavailable) and 25 of 71 genes (4 unavailable) identified in the UKB combined model analysis also reached significance in the replication analysis ( **Fig. S4**, **Tables S11–12**). The top SNPs in the RS, rs2290911 (chr2) and rs1800407 (chr15), were likewise the strongest signals in the UKB. Similarly, the top three genes, *ACP1*, *ALKAL2*/*FAM150B*, and *SH3YL1*, were identical across both cohorts and remained genome-wide significant in the RS. The remaining signals showed consistent directions of effect (see **Fig. S4c**), with enrichment beyond chance even when not significant, supporting robust genetic replication of RAG in the independent RS cohort.

### Saliency and attention maps highlight the optic disc, macular area, and blood vessels

To better understand what the models rely on to predict age, we first computed saliency maps from the fine-tuned models using RELPROP [29], as in the original RETFound publication [25]. Saliency maps showed the strongest activation in the macula and the temporal half of the optic disc, with sex-specific models displaying patterns highly similar to those of the combined-sample models (**Fig. 5a**, top row). A comparison between the youngest and oldest age quartiles revealed distinct patterns: in younger retinas, saliency was concentrated primarily in the macula ( **Fig. 5b**, top row), whereas in older retinas, both the macula and the temporal optic disc contributed to the model’s predictions (**Fig. 5c**, top row). Saliency maps were highly consistent across model folds and showed strong symmetry between left and right eyes (**Fig. S5**).

**Figure 5.**
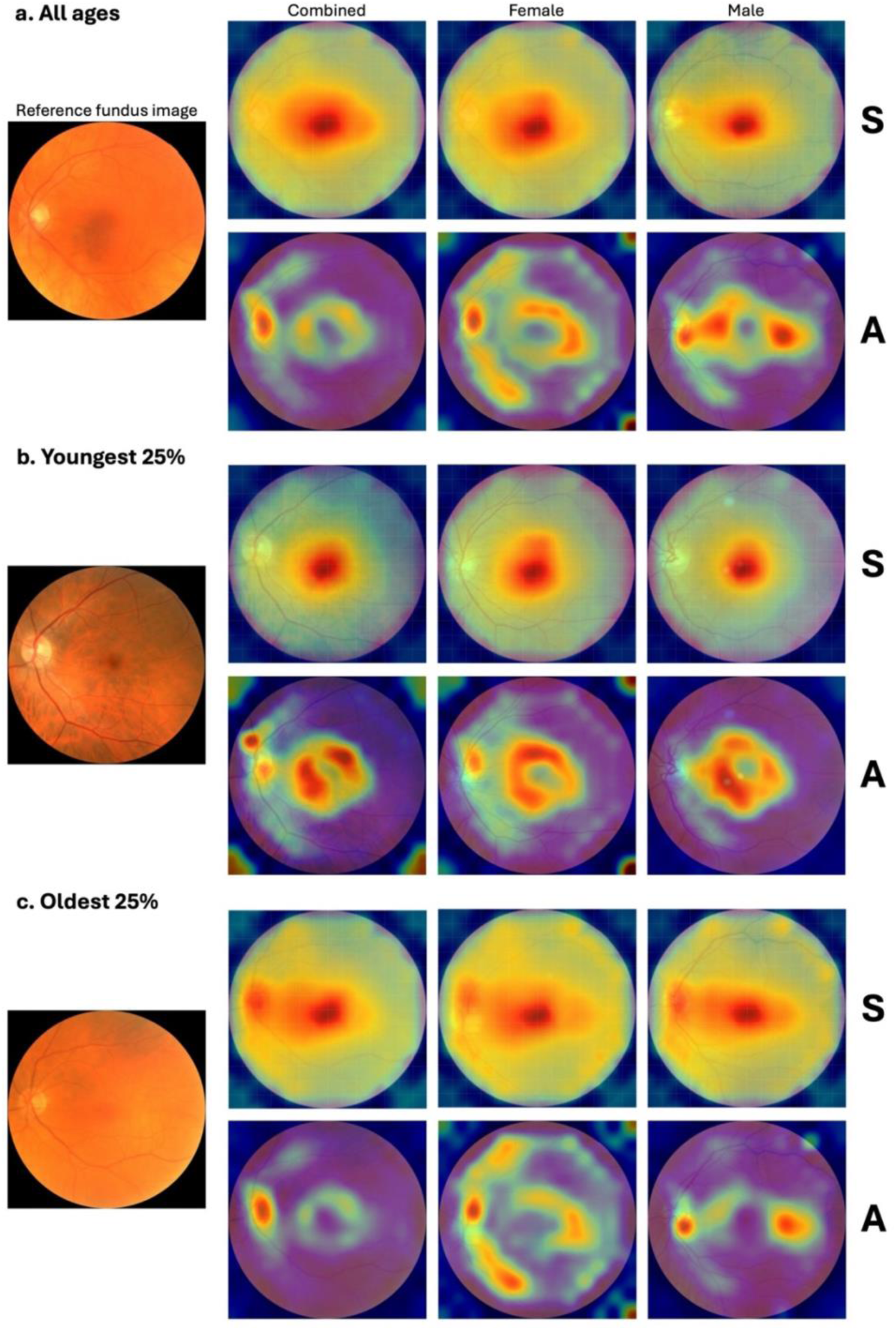
Group-average saliency and attention maps of the fine-tuned models on left eye images. Smoothed pixel-level saliency (S, upper rows) and attention (A, lower rows) maps of fold 1 models, obtained by bicubic interpolation from patch-level maps, are projected on a reference fundus image for **(a)** all ages (n=13,078 images combined sample, 5897 females and 5838 males), **(b)** the youngest quartile (n=3270 combined, 1475 females and 1460 males), and **(c)** the oldest quartile (n=3269 combined, 1474 females and 1460 males).

Attention maps extracted from attention blocks of the Vision Transformer were partially consistent with the saliency maps, highlighting the temporal optic disc across all groups, especially in the oldest quartile. However, unlike the saliency maps, attention maps emphasized the area surrounding the macula rather than the macula itself (**Fig. 5**, bottom rows). Additionally, the main vascular arches emerged as prominent regions of attention, particularly in female participants in the oldest quartile (**Fig. 5**, middle column, bottom row), suggesting that the model may rely more heavily on vascular features when predicting age in older female retinas. As observed with saliency maps, attention maps exhibited strong left–right eye symmetry, although they showed greater variability across model folds (see **Fig. S6**). Detailed maps from the 16 individual attention heads of the last block are shown in **Figures S7–S9**.

### Model latent variables reflect vascular density, diameter, and bifurcations

We then examined the correlations between the model’s latent variables (LVs) and interpretable image features, including vascular morphology, optic disc area, and RGB statistics. Most LVs were correlated with vascular density and bifurcation count (mean |*r*| ∈ [0.26, 0.34]). Notable correlations were also observed with central retinal arterial and venous equivalents (mean |*r|* = 0.17 and 0.13), artery diameter SD (mean |*r*| = 0.14), and to a lesser extent, median vascular diameters, and green channel mean and SD (mean |*r*| ∈ [0.06, 0.14]) (**Fig. 6**). The correlation pattern was highly consistent across all 15 models (5 folds × 3 groups), with correlation matrices showing strong agreement across folds and sex groups (**Figs. S10–S23**).

**Figure 6.**
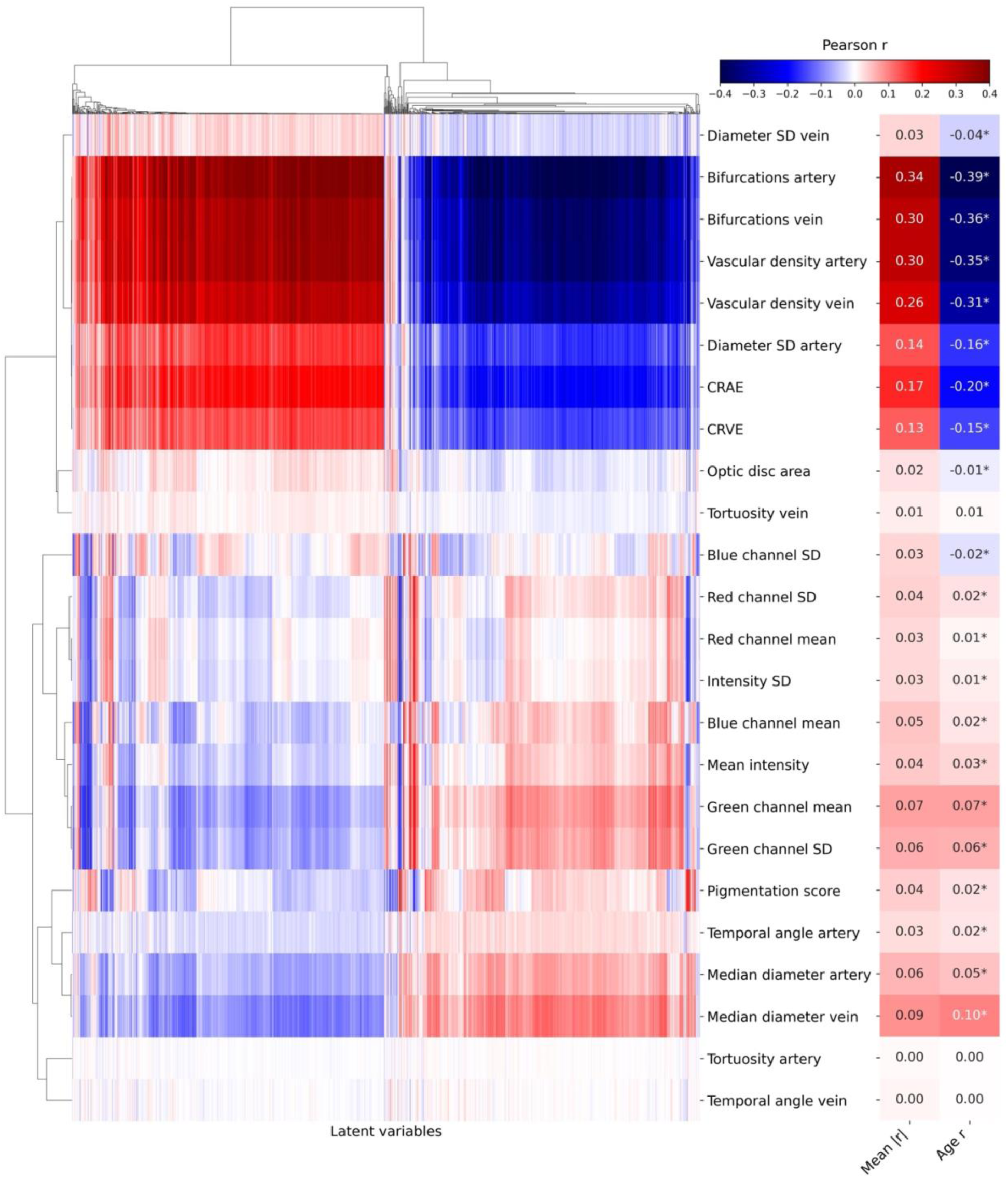
Correlation map between the model’s 1024 latent variables and interpretable image features, in the combined sample for fold 1 (n=24,044 images). For every image feature, the mean absolute correlation with latent variables and the correlation with chronological age are shown on the right side of the figure. CRAE: central retinal arterial equivalent, CRVE: central retinal venous equivalent, SD: standard deviation, *: *p*<0.05.

Interestingly, the correlation between image features and chronological age (shown for each feature in the rightmost column of **Fig. 6**) closely mirrored their correlation with the LVs. Vascular features such as density and bifurcations showed stronger correlations with age than with the LVs, while the opposite pattern was seen for RGB-based features, which correlated more with LVs than with age. This suggests that although the model’s internal representations reflect vascular aging to some extent, they also encode additional non-age-related variation, particularly in color distribution and intensity, that may contribute to its age estimates.

## Discussion

In this study, we fine-tuned RETFound to predict chronological age from retinal images, demonstrating robust generalization to an independent dataset with improved accuracy through averaging predictions from both eyes. We trained sex-specific models and found differences in RAG and its associations with clinical outcomes and genetic variants between females and males. RAG was associated with key health outcomes including CVD, cancer, and death, as well as eye disease, metabolic and inflammatory biomarkers. Genetic analyses identified novel and previously reported associations involved in aging-relevant pathways. Attention maps and latent space analyses pointed to the temporal optic disc, macular area, and blood vessels as important regions, and revealed correlations with vascular features.

With an MAE of 2.85 years and an R^2^ of 0.80, RETFound fine-tuned to predict age on UKB data outperformed previous CNN-based models, which have reported MAE between 3.08 and 4.5 years and R^2^ between 0.51 and 0.77 [3,6,7,9,10]. Although foundation models require considerable computational resources to pretrain, our results suggest they offer a measurable benefit in downstream regression tasks such as age prediction. When applied to the independent RS dataset, the model maintained a high correlation between predicted and true age but exhibited a consistent downward bias, especially in males. This systematic underestimation may be partially explained by the slightly older age distribution in the RS cohort (on average 0.3 years older than the UKB), combined with regression-to-the-mean effects that tend to produce larger underestimations at the upper end of the age range prior to bias correction. Furthermore, the lower age predictions in the RS could be due to technical differences between datasets, such as the camera model or image acquisition settings, or it may reflect genuine differences in participants’ health, whereby RS retinas may appear biologically younger than UKB retinas.

Consistent with previous findings [5,9], we show that averaging predictions across images from both eyes improves accuracy, likely by reducing noise arising from variations in image quality or other image parameters. While this strategy may not be ideal in clinical datasets, as it may obscure pathologies affecting only one eye, or with asymmetrical disease progression, it is well-suited to address image quality variability in population-level datasets like the UKB. Therefore, the choice to average or not the predictions coming from both eyes should be guided by the specific research question and context of a given study.

We observed a modest but consistent sex difference in predicted retinal age across cohorts, with male retinas estimated as appearing younger. This stands in contrast to brain age models, which often estimate younger brain age in females [17–19], although findings vary across studies [20–22]. Both brain and retinal age predictions may be influenced by overall body or organ size. Brain age studies typically account for intracranial volume by using corrected image-derived phenotypes, whereas in our study, age was predicted directly from raw retinal images without adjusting for potential effects of eye size. Given that females and older individuals tend to have smaller bodies and possibly eye sizes on average, we hypothesize that this may have confounded the model, contributing to the observed overestimation of age in females. Supporting this, the sex difference in RAG was attenuated, though not eliminated, after adjusting for height. Further research is needed to explore anatomical scaling, physiological, hormonal or metabolic factors as potential drivers of the observed sex difference in retinal age predictions.

Interestingly, while cross-sectional associations between RAG and health indicators were generally stronger in males, RAG predicted incident CVD similarly in males and females, after adjusting for conventional risk factors such as BMI, smoking, SBP, cholesterol, and diabetes. In fact, HRs were higher in females for stroke, thrombosis, and overall CVD—though not significantly so, likely due to limited statistical power (**Table S4**). While males have a higher overall risk of CVD [29], our findings suggest that this difference may be largely captured by traditional risk factors included in the models. In contrast, the additional variance captured by RAG may be more informative in females, potentially reflecting sex-specific biological aging processes not well represented by traditional risk metrics. For example, women exhibit higher baseline inflammation, including elevated CRP and interleukin-6, which has been linked to greater CVD risk compared to men, even after adjusting for risk factors like smoking and BMI [30]. Hormonal dynamics and sex-specific metabolic changes in females may contribute to patterns of aging that are not well captured by conventional risk metrics, but are more readily reflected in RAG.

Conversely, RAG predicted future cancer and dementia risk in males but not in females. The association with cancer may reflect known biological differences: male cells experience greater oxidative stress and accumulate more somatic mutations than female cells [31], both of which increase susceptibility to malignant transformation and represent hallmarks of aging that may be captured by RAG. For dementia, we pooled all dementia subtypes due to limited case numbers, but we hypothesize the observed association may have been driven primarily by vascular dementia, given its shared link with metabolic syndrome and cardiovascular risk factors [32].

Genetic analyses of RAG revealed both novel loci and replication of previously reported associations [9,10,33]. Novel loci had been previously associated with corneal resistance factor (rs1200105) [34], educational attainment (rs79073127 and rs6477724) [35], and skin and eye cancer (rs12203592 and rs2413887) [36,37]. At the gene level, unlike traditional approaches that link only the most significant SNP to the nearest gene, our method aggregates signals across all SNPs within each gene region while accounting for LD, thereby improving sensitivity to detect biologically meaningful associations [28]. The genes we identified have been implicated in metabolism, lifespan, and retinal vessel density (*ACP1* [38,39], *SH3YL1* [39–41], *ALKAL2* [39,42], *MACF1-BMP8A* [43]), in age-related eye diseases and retinal vessel diameter variability (*GJA3* [44], *ARMS2* [45], *NPLOC4*-*PDE6G* [39,46]), cellular senescence (*UBN1*) [47], Alzheimer’s disease (*CCDC25-ESCO2*) [48], and immune function (*PTPN7*) [49] among others. This is consistent with the clinical traits linked to RAG, supporting its biological relevance. Like previous retinal age studies [9,10,33], we also identified *OCA2*, a gene involved in eye pigmentation, suggesting that image features that are unlikely to be age-relevant may influence the model prediction. Among the female-specific associations, we identified two genes previously linked to retinal vessel tortuosity (*CAPN12* and *ACTN4*) [39,50,51], in line with our observation that model attention maps highlighted blood vessels more prominently in females.

Saliency maps consistently showed prominent model activation in the macula and temporal half of the optic disc, while attention maps partially aligned by highlighting the optic disc and surrounding macular regions. Crucially, attention maps also emphasized the main vascular arches in females—particularly in older retinas—mirroring our latent-variable analysis, which showed strong correlations with vessel density, bifurcation count, and vessel diameter. We hypothesize that the number of visible small vessels, which typically decreases with age [52], plays a role in model predictions. Interestingly, a recent study reported higher macular vascular density in males compared to females, with sex differences varying between foveal and parafoveal areas and across age groups [53]. This anatomical variation could help explain our observation that saliency maps focused on the fovea while attention maps emphasized the surrounding macular region. These findings suggest that the parafoveal vasculature may carry both age-relevant and sex-discriminative information, contributing to the model’s ability to capture distinct biological aging patterns between sexes.

Finally, our findings must be interpreted within the context of the study’s limitations. Although RETFound was pre-trained on data with some ethnic diversity, both the UKB and RS cohorts analyzed here were predominantly composed of individuals of White European ancestry. As such, the generalizability of our results to other populations remains to be evaluated. Broader representation in training datasets will be essential to ensuring equitable performance across ancestries. Furthermore, both cohorts represent relatively healthy, aging populations; as such, the findings may not generalize to clinical populations with higher disease burden or atypical ocular presentations. The limited number of incident dementia cases, particularly within sex-specific strata, restricted our ability to examine subtype-specific associations with sufficient statistical power. Finally, despite leveraging saliency and attention maps, as well as correlations with interpretable image features, the precise biological mechanisms underlying the age-related signals captured by the models remain incompletely understood.

Overall, our study demonstrates that RAG offers a non-invasive, scalable biomarker that complements traditional indicators of biological aging. While training sex-specific models did not improve predictive accuracy, the distinct genetic and clinical correlates observed between females and males underscore the value of sex-stratified analyses in aging research. Incorporating sex-specific perspectives will therefore be essential for refining aging biomarkers and advancing their application in precision medicine.

## Methods

### Imaging data

We used color fundus images (CFIs) from UK Biobank (UKB) instances 0 and 1. These single-field, 45° color photographs, centered to capture the optic disc and macula, were acquired using a Topcon 3D OCT-1000 Mark II digital ophthalmic camera. To ensure image quality, we applied a quality control (QC) procedure based on the method described by Zekavat et al. [41], excluding the lowest-quality 25% of images. Briefly, a quality score was assigned to each image based on a deep learning model trained to identify poor-quality scans, and images below the 25th percentile of the score distribution were removed. This resulted in a final dataset of 130,360 images from 71,343 participants. We cropped black borders from all retained images [54]. Participants were between 39.2 and 79.1 years old (mean age = 57.7 ± 8.2 years). For the sex-specific analyses, we created age-matched cohorts of males and females, each comprising 32,918 individuals with a mean age of 57.9 ± 8.3 years and an age range of 40.1 to 75.8 years.

For the external validation of our models, we used data from the Rotterdam Study (RS), a prospective, population-based cohort study conducted in Ommoord, a district of the city of Rotterdam, The Netherlands [55]. It comprises four cohorts, initiated in 1989, 2000, 2006, and 2015, with a total of 17,931 participants. Participants undergo examinations approximately every five years, which include retinal imaging using various imaging modalities and camera systems across study rounds. For the model replication, CFIs acquired with the Topcon 3D OCT-2000 FA plus camera with a macula-centered field-of-view were used, as this modality was deemed most comparable to the UKB imaging while offering broad availability within the RS. In total, 37,771 fundus images were available from 7480 participants (14,912 eyes across 8813 visits). After filtering by age (40 < age ≤ 70 years, a range encompassing 97% of the UKB sample) and image quality (excluding the lowest 25%), 10,354 images from 4757 participants were retained for analysis (mean age = 58.0 ± 7.2 years). For the genetic analysis replication, we aimed to maximize sample size by selecting one assessment per participant according to the following rules: preferentially retaining Topcon 3D OCT-2000 FA macula-centered images when available, and otherwise using the closest available modality. When multiple assessments were present, we chose the visit with the highest average image quality. We applied the same age and quality filters. This resulted in a dataset of 11,368 participants (mean age = 60.6 ± 6.4 years) with 88,789 fundus images.

### RETFound fine-tuning

RETFound is a Vision Transformer-based foundation model pre-trained on a large collection of unlabeled retinal images using self-supervised learning [25]. Originally designed for classification tasks, we adapted the architecture for regression to predict chronological age from CFIs. We employed a five-fold cross-validation framework, where participants were randomly divided into five non-overlapping groups. For each fold, we trained a separate model using 60% of participants for training, 20% for validation, and 20% for testing, with training, validation, and test sets rotating across folds. We used all available images, ensuring that, for a given fold, images from the same participant were assigned to the same set to avoid leakage between sets.

We resized images to 224 × 224 pixels. For data augmentation during training, we applied random crops and rescaling with a scale range of (0.8, 1.0) and aspect ratio range of (0.9, 1.1), along with random rotations up to 20°. We normalized all images using the ImageNet RGB mean and SD.

We trained models for 50 epochs, including 10 warm-up epochs. The learning rate followed a half-cycle cosine decay schedule: it increased linearly during the warm-up phase, then decreased following a cosine curve toward a minimum value. Specifically, the learning rate was initialized as 0.005×batch size/256, and decayed to a minimum of 1×10^−6^. We used mean absolute error (MAE) as the loss function. The batch size for training was 16. Additional training hyperparameters included a drop path rate of 0.2, weight decay of 0.05, and layer-wise learning rate decay of 0.65. With the exception of the adjustments required for regression, we maintained the original RETFound training configuration as closely as possible.

We repeated the same five-fold cross-validation procedure separately for female and male participants using sex-specific, age-matched samples. This resulted in a total of 15 models: five for the combined sample, five for females, and five for males. This cross-validation setup ensured that each image had an out-of-sample prediction, allowing for robust performance assessment across the entire dataset. We evaluated the model performance using the MAE, Pearson’s correlation (*r*) and coefficient of determination (R^2^) between the predictions and the chronological ages. For external validation in the RS, we applied the five combined-model weights to the entire RS dataset and averaged the predicted ages to produce a single ensemble prediction per image. To evaluate overall performance, we computed the MAE, *r*, and R^2^ between the ensemble predictions and the true ages. To assess the variability across individual models, we calculated the SD of the five model predictions for each image and reported the mean of these per-image SDs across the dataset. We repeated this procedure using the sex-specific models on the corresponding female and male RS samples.

### Retinal age gap

To correct for regression-to-the-mean bias in predicted age, we applied a post hoc adjustment following the procedure described by Cole et al. [18]. Specifically, we fit a linear regression of predicted age on chronological age to estimate the slope (𝛼) and intercept (𝛽), then adjusted predicted age by subtracting 𝛽 and dividing by 𝛼.

We then defined the retinal age gap (RAG) as the difference between the corrected predicted age and the participant’s chronological age. To assess sex differences, we compared mean RAG values between females and males using independent-sample t-tests and quantified the effect size with Cohen’s *d*. We performed this analysis in both the UKB and RS datasets, while subsequent disease associations were conducted using UKB data only.

### Association with health and lifestyle factors

To investigate the relationship between RAG and various health outcomes, we conducted a cross-sectional analysis using data from the UKB. For participants with imaging data available at multiple time points, we used the first available instance.

Continuous outcomes included body mass index (BMI; data-field (DF) 21001), waist-to-hip ratio (WHR) defined as waist circumference (DF 48) divided by hip circumference (DF 49), systolic blood pressure (SBP; DF 4080 averaged across measurements), diastolic blood pressure (DBP; DF 4079 averaged across measurements), log-transformed pack-years of smoking (DF 20161), glycated hemoglobin (HbA1c; DF 30750), glucose (DF 30740), white blood cell (leukocyte) count (DF 30000), C-reactive protein (CRP; 30710), neutrophil (DF 30140) to lymphocyte (DF 30120) ratio (NLR), insulin-like growth factor 1 (IGF-1; DF 30770), low-density lipoprotein (LDL) cholesterol (DF 30780), high-density lipoprotein (HDL) cholesterol (DF 30760), triglycerides (DF 30870), total cholesterol (DF 30690), the VO₂max cardiorespiratory fitness estimate (DF 30038), sleep duration (DF 1160), reasoning score (also called fluid intelligence score; DF 20016), reaction time (DF 20023), and log-transformed telomere length (DF 22192). For participants who took antihypertensive medication, we added 10 mmHg to SBP and 5 mmHg to DBP values. Similarly, for participants who took cholesterol-lowering medication, we added 1.4 mmol/L to LDL, 0.4 mmol/L to triglycerides, 1.6 mmol/L to total cholesterol, and subtracted 0.1 mmol/L from HDL. Sex-specific associations additionally included testosterone (DF 30850) for males, and age at menopause (DF 3581) for females.

Binary outcomes included the use of diabetes, cholesterol, or BP medication (DFs 6153 and 6177), diabetes—defined as glucose >= 11.1 and/or HbA1c >= 48 and/or self-reported diagnosis by a doctor (based on DFs 2443 and 2976) and/or diabetes medication use—, hypertension— SBP > 140 and/or DBP > 90 and/or BP medication use—, dyslipidemia—cholesterol >= 5 and/or HDL <= 1 and/or LDL >=4.1 and/or triglycerides >= 2.2 and/or cholesterol medication use—, eye diseases (DF 6148) including cataract, glaucoma, and macular degeneration, cataract surgery (DF 5324), cancer history (DF 2453), metabolic syndrome—defined according to the International Diabetes Federation consensus [56] as having three or more of the following: large waist circumference (>= 80 cm for female or >= 94 cm for males), triglycerides > 1.7 or cholesterol medication, low HDL (< 1.29 for females or < 1.03 for males), high BP (SBP >= 130 and/or DBP >=85 and/or BP medication), and high glucose which we substituted with the above definition of diabetes since fasting glucose was not available—, smoking status (current/previous vs. never; DF 20116), alcohol intake (occasional/regular vs. never; DF 1558), having children (binarized from DFs 2734 and 2405), visual memory (>= median in the pairs matching task; DF 399), and prospective memory (correct recall in first attempt; DF 20018). Additionally for females, we included the menopausal status (DF 2724), contraceptive pill and hormone replacement therapy (HRT; DF 6153).

All models controlled for age (derived from DFs 34, 52, and 53), age-squared, sex (DF 31), height (DF 50), height-squared, self-reported ethnic background (DF 21000, binarized as White/British vs. other given the low number of non-White participants), the first 10 genetic principal components (PCs; DF 22009), educational attainment (DF 6138)—categorized as low (“CSEs or equivalent”, “NVQ or HND or HNC or equivalent”, “Other professional qualifications eg: nursing, teaching”, or “None of the above”), medium (“A levels/AS levels or equivalent”, “O levels/GCSEs or equivalent”), or high (“College or University degree”) [57]—, household income (DF 738)— categorized as low (< 18,000 £), medium (between 18,000 and 51,999 £), or high (>= 52,000 £)—, and Townsend Deprivation Index (DF 22189). We imputed missing covariate data using the group mean (for continuous covariates) or mode (for categorical covariates).

We standardized (z-scored) continuous outcomes and covariates, and excluded values that lay beyond 4 SD from the group mean. We applied linear regression models for standardized continuous outcomes and logistic regression for binary traits, with RAG entered as a standardized predictor. We conducted sex-stratified analyses to explore differential associations, and reported effect sizes as standardized beta coefficients or odds ratios. We corrected p-values for multiple testing (122 tests) using false discovery rate (FDR) [58] with an alpha threshold of 0.05.

### Survival analysis

To assess the prospective associations between RAG and major health outcomes, we conducted Cox proportional hazards regression analyses using follow-up data spanning approximately 15 years from the UKB. We performed time-to-event analyses for all-cause mortality (DF 40000) and incident cases of chronic obstructive pulmonary disease (DFs 42016, 131486, 131488, 131490, and 131492), cancer (DF 40005), dementia (DFs 42018, 130836, 130838, 130840, and 130842), and cardiovascular disease (CVD), which encompassed stroke (DFs 42006, 131366, 131362, and 131364), thrombosis (DFs 131308, 131388, and 131400), myocardial infarction (DFs 131298, 131300, 131302, and 42000), ischemic heart disease (DFs 131296, 131304, and 131306), and non-ischemic cardiomyopathy (DFs 131338, 131340, 131288, and 131292). We adjusted models for age, sex, height, ethnicity, education, income, Townsend, SBP, HDL, total cholesterol, smoking status, BMI, diabetes, and BP medication. We conducted analyses in the full sample and stratified by sex to examine potential sex-specific associations. We corrected p-values for multiple testing (30 tests) using FDR [58] with an alpha threshold of 0.05.

### Genome-wide association study

We performed a genome-wide association study (GWAS) of RAG using Regenie [59], which accounts for relatedness and population structure via a two-step ridge regression approach. Before conducting the analyses, we applied filters for minor allele frequency (MAF ≥ 0.01), minor allele count (MAC ≥ 100), genotype missingness (≤ 0.1), and Hardy–Weinberg equilibrium (HWE, p ≥ 1×10⁻¹⁵). Because quality control of the X chromosome remains debated given its biological specificities, we implemented additional steps [60]. We handled the X chromosome separately in each sex, using a relaxed missingness threshold for males (≤ 0.2) and excluding all ampliconic regions. We did not apply HWE filtering to the X chromosome, as the HWE test does not accommodate hemizygous males and may otherwise eliminate valid variants. Prior to the analysis, we applied a rank-inverse normal transformation to the RAG phenotype. The covariates included in the GWAS were age, sex (DF 31)—excluding cases of aneuploidy (DF 22019) or when genetic sex (DF 22001) did not match recorded or self-reported sex (DF 31)—, age-squared, age-by-sex, age-squared-by-sex, height, height-squared, instance, assessment centre (DF 21003), genotype measurement batch (DF 22000), spherical power (DFs 5084 and 5085), spherical power-squared, cylindrical power (DFs 5086 and 5087), cylindrical power-squared, and the first 20 genetic PCs. For sex-specific GWAS, the covariates were the same excluding sex.

We computed gene-level association scores using PascalX [28], which aggregates single nucleotide polymorphism (SNP)-level statistics into gene scores using the approximate “saddle” method. We scored protein-coding genes, including all SNPs with MAF ≥ 0.05, located within a 50 kb window around each gene. We used the UK10K reference panel [61] for gene scoring on the autosomes while we built the X chromosomal panel independently for females and males using 5000 unrelated white individuals per sex from the UKB.

We estimated SNP-based heritability (h^2^_SNP_) of RAG using the genomic-relatedness-based restricted maximum-likelihood (GREML) method implemented in the GCTA (Genome-wide complex trait analysis) software package [62]. We fitted a genetic relationship matrix (GRM) constructed from common (MAF > 0.01) and directly genotyped variants to estimate the proportion of variance explained in RAG by all measured SNPs.

For the replication analysis, because imputation and QC were conducted independently in the four RS sub-cohorts, we first ran GWAS within each cohort and subsequently combined the results in a meta-analysis. For the individual GWAS, we applied linear regression models in PLINK 2.0 [63], adjusting for age, sex, age-squared, age-by-sex, age-squared-by-sex, height, height-squared, spherical power, spherical power-squared, cylindrical power, cylindrical power-squared, and the first 10 genetic PCs. We then meta-analyzed results across cohorts using an inverse-variance weighted fixed-effects approach implemented in METAL [64]. We derived association p-values from the corresponding *z*-statistics. We computed gene scores using PascalX [28] with the parameters described above. To assess replication of genetic findings, we adopted a candidate approach, applying FDR correction with a threshold 0.05 to p-values from SNPs, respectively genes, that were genome-wide significant in the UKB. We also computed Pearson’s correlation between SNPs effect sizes (*β*) in the UKB and in the RS to assess consistency of results regardless of significance.

### Saliency and attention maps

To interpret the internal representations learned by RETFound and visualize the model’s implicit focus during prediction, we applied two distinct methods.

First, we generated saliency maps using RELPROP [29], a method that applies layer-wise relevance propagation (LRP) tailored for Transformer-based architectures, as used in the original RETFound publication [25]. RELPROP reveals model focus by integrating relevance scores with gradient information and propagating relevance backward through the whole network while preserving the total relevance across layers, creating more accurate attribution than conventional attention visualization alone. To accurately capture RETFound’s prediction process, we adapted the RELPROP method to handle the global pooling architecture by distributing relevance equally across all non-CLS tokens, ensuring proper attribution for the regression pathways. Furthermore, we generated group-average saliency visualizations that highlight image regions consistently attended to by the model across a large set of input images, revealing which anatomical structures drive the model’s decision-making process at the population level. This approach is meaningful in our context, as we restricted the analysis to left eye images (right eye saliency maps are shown in **Fig. S4**), ensuring that key anatomical features such as the optic disc and macula were consistently located across individuals.

Second, we extracted attention maps from the 24 attention blocks of the Vision Transformer, and computed attention rollout [65]. Specifically, we captured the spatial attention scores of the class token directed towards the patch tokens (“CLS→patch attention”), a commonly used proxy for identifying important image regions in Transformer-based models [66]. We implemented a forward hook to extract the softmax-normalized attention weights from each block. These weights represent the flow of information within the Transformer, indicating how much the CLS token relies on other patches, and form 14×14 spatial attention maps for each of the 16 attention heads. We then averaged the attention across all heads and aggregated across all blocks to obtain a single attention map per image [65]. Finally, similarly to saliency maps, we averaged attention maps across the cohorts to produce population-level visualizations of the regions most consistently attended to by the model.

Saliency maps and attention maps represent different but complementary approaches to interpreting Transformer-based models. Saliency maps operate in a backward direction, tracing the model’s decision back through the network using relevance scores combined with gradients. This yields causal attributions by accounting for non-linear interactions across all layers and attention heads. In contrast, attention maps are obtained in the forward pass, using the softmax-normalized weights from the attention layers. These reflect the model’s internal information routing (i.e., how much each input token is attended to by the CLS token), but do not indicate actual influence on the output, making them associative rather than causal. Because saliency maps trace decision-making and attention maps reveal focus patterns, differences between them can be informative. For example, the model might consistently attend to certain patches for context, while relying on different regions to make the final prediction.

### Linking latent representations to interpretable image features

To investigate which interpretable image features the models may rely on to predict age, we examined Pearson’s correlations between the 1024 latent variables (LVs), derived from the encoder portion of the models, and a comprehensive set of handcrafted retinal features. Specifically, we computed correlations between each LV and 24 image-derived variables: mean image intensity defined as the mean of the red, green, and blue channels, SD of image intensity, mean and SD of the red, green, and blue channels separately, optic disc area, central retinal arterial equivalent, central retinal venous equivalent, median and SD of artery and vein diameters, artery and vein tortuosity, artery and vein bifurcation counts, artery and vein temporal angles, artery and vein vascular density, and pigmentation score. All vascular features were extracted using the VascX software [67]. For the optic disc segmentation, we annotated a random set of 100 CFIs and used them to re-train a publicly available U-Net model [68]. We computed the pigmentation score using the pipeline published in [69], which was developed to decouple traditional demographic variables from clinical imaging characteristics and has been previously validated on the UKB data. We computed correlations separately for each of the 15 trained models (across five cross-validation folds and three sex groups). Additionally, for the 24 image features, we computed correlations with the chronological age.

### Data and code availability

UK Biobank data are available upon successful application (https://www.ukbiobank.ac.uk/enable-your-research/apply-for-access). RETFound code is publicly available at https://github.com/rmaphoh/RETFound. The code used to generate the presented results will be made available on GitHub upon publication (https://github.com/ot710/retinal-age).

## Data Availability

UK Biobank data are available upon successful application (https://www.ukbiobank.ac.uk/enable-your-research/apply-for-access). The code used to generate the presented results will be made available on GitHub upon publication (https://github.com/ot710/retinal-age).

## Acknowledgements

This research has been conducted using the UK Biobank Resource under Application Number 90947. This work was funded by the Swiss National Science Foundation grant no. CRSII5 209510 for the “VascX” Sinergia project.

