## Supplementary Figures for "Deep learning aging marker from retinal images unveils sex-specific clinical and genetic signatures"

**a**

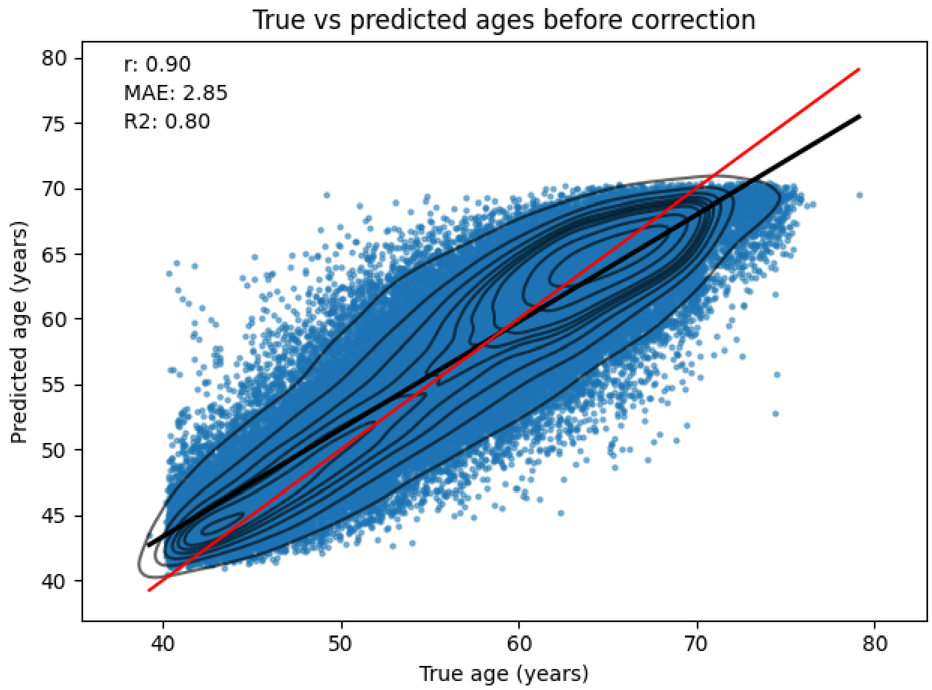


**b**

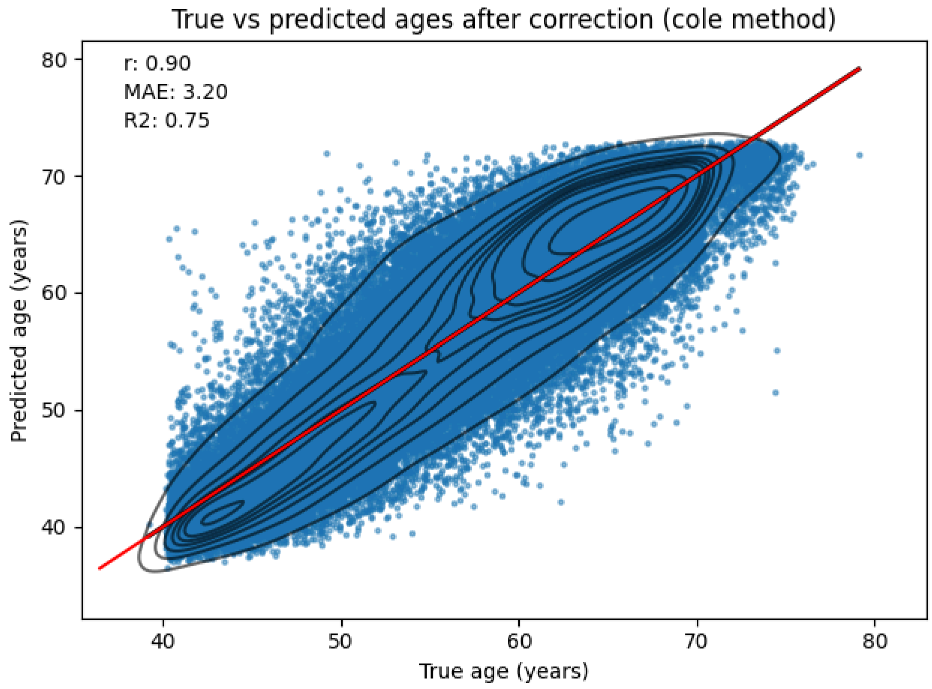


**Figure S1.** Predicted age against chronological (true) age **(a)** before and **(b)** after correction for the regression-to-the-mean bias, in the combined sexes model in the UK Biobank. The red line is the diagonal, while the black line is the linear regression fit line (both lines are superimposed in **b**). r: Pearson's correlation, MAE: mean absolute error, R2: coefficient of determination.

**a**

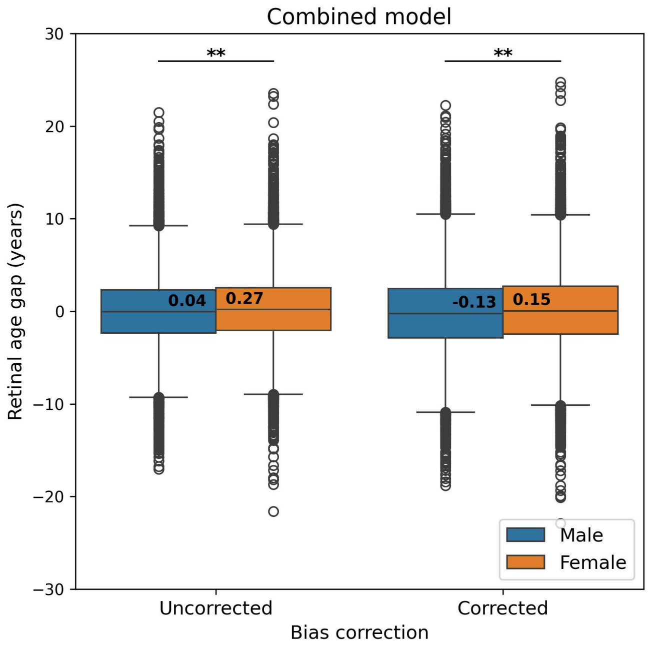


**b**

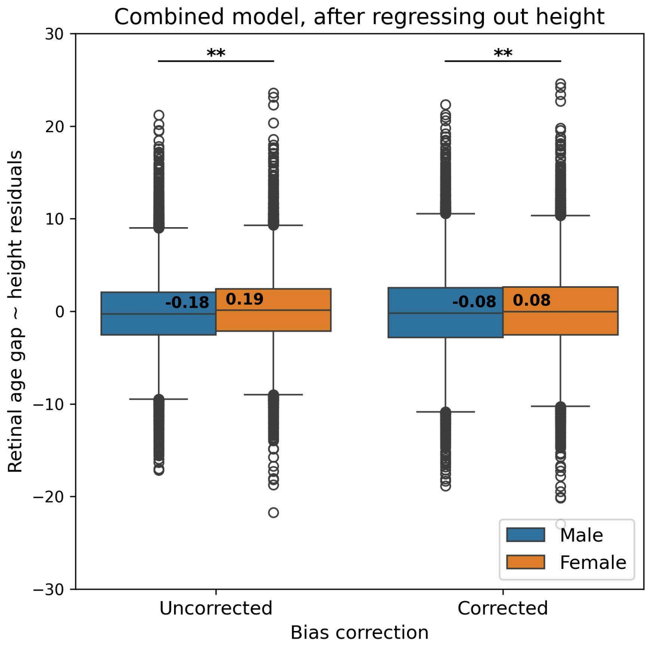


**Figure S2.** Sex comparison of retinal age gap from the combined-sample model, with and without correction for the regression-to-the-mean bias, **(a)** before and **(b)** after regressing out height from the retinal age gap. **: *p*<0.001 from t-test comparing sexes.

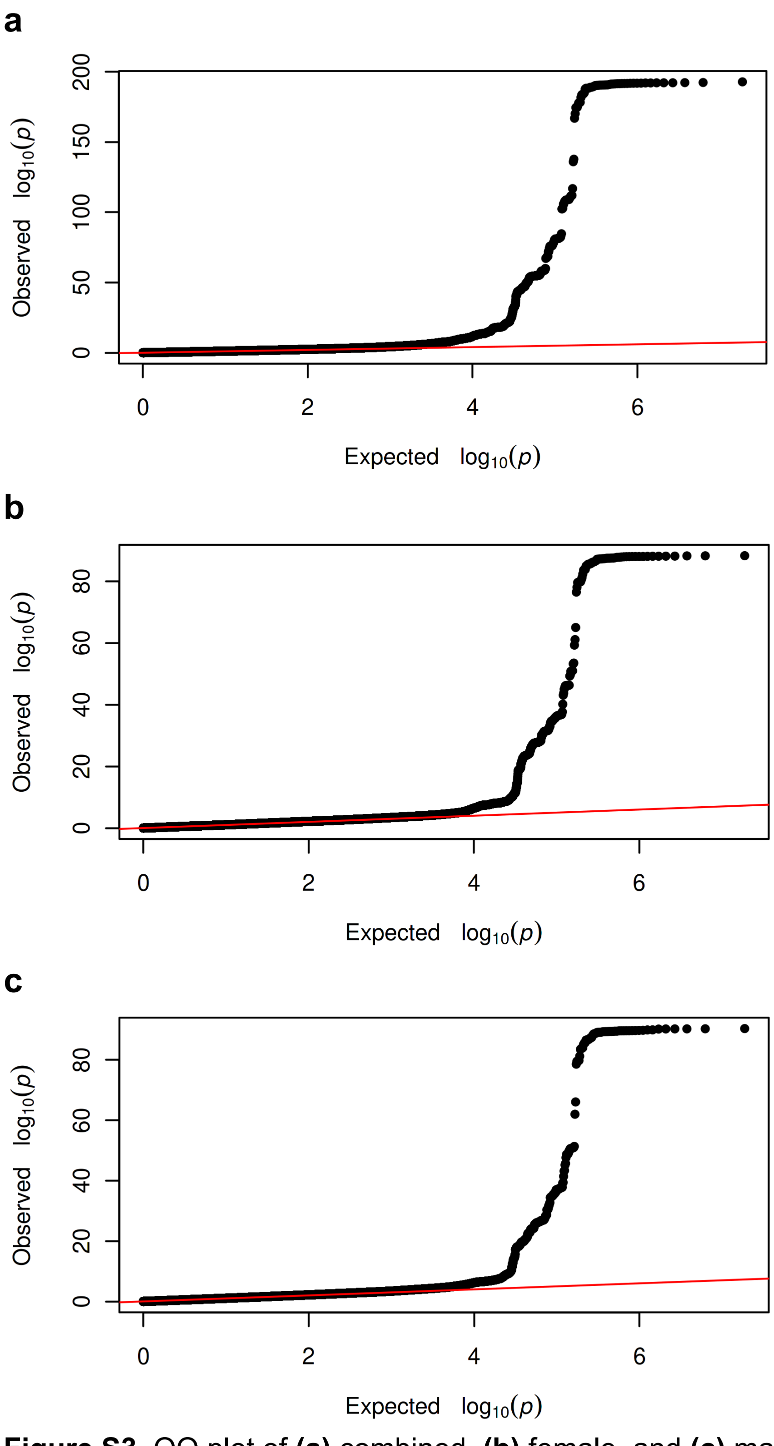


**Figure S3.** QQ plot of **(a)** combined, **(b)** female, and **(c)** male retinal age gap GWAS.

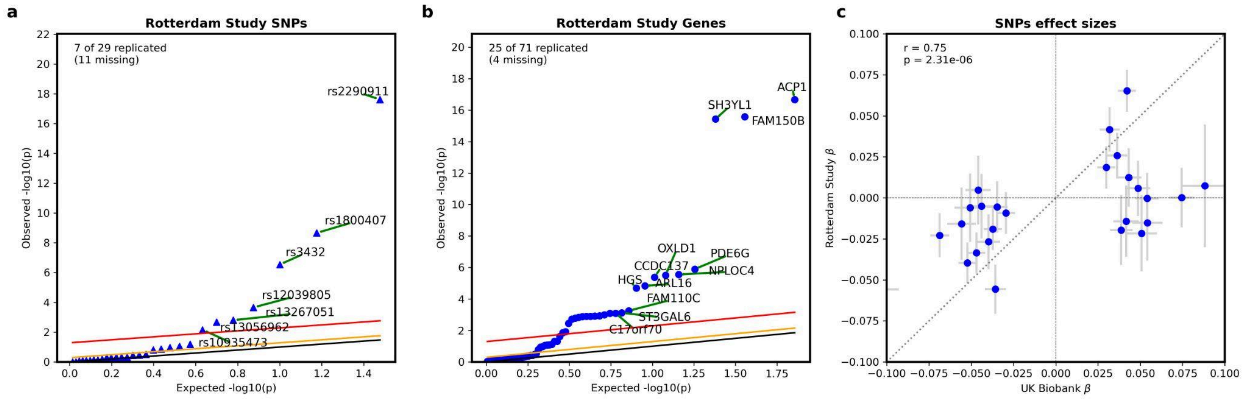


**Figure S4. Replication of genetic analysis in the Rotterdam Study (n=9547).** Replication of **(a)** variants (SNPs) and **(b)** genes that were significant in the UK Biobank. The red line represents a false discovery rate (FDR) of 0.05, the orange line represents a FDR of 0.5, and the black line represents observed = expected. The label "missing" indicates that those SNPs or genes were significant in the UK Biobank, but not available in the Rotterdam Study. **(c)** Effect sizes (betas) of significant SNPs in the UK Biobank against their effect sizes in the Rotterdam Study. Error bars represent standard errors. Pearson's correlation between the two cohorts was 0.75 ($p$<0.0001).

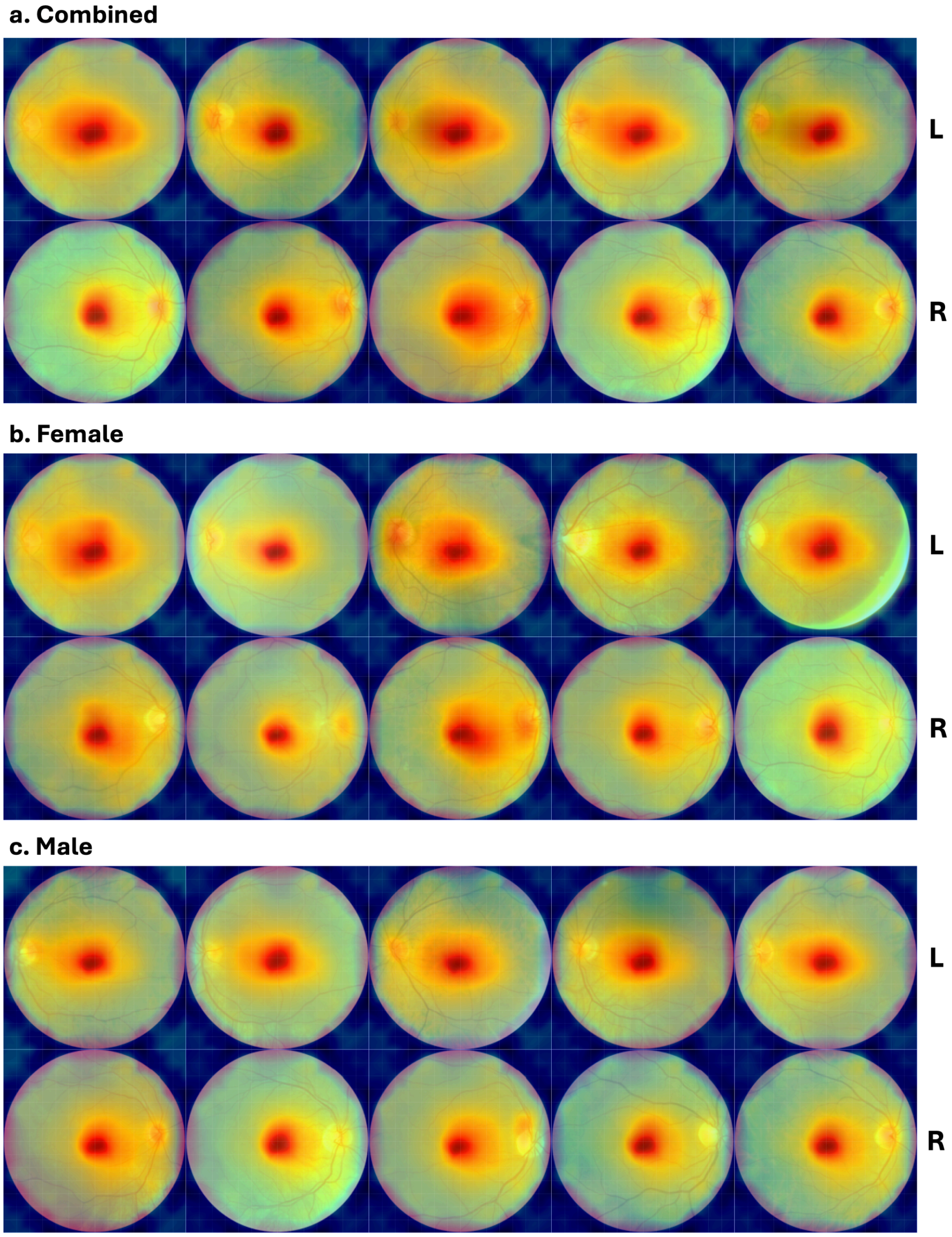


**Figure S5. Group-average saliency maps in the 5 folds of the fine-tuned models.** Smoothed pixel-level saliency maps, obtained by bicubic interpolation from patch-level maps, are projected on a reference fundus image for **(a)** the combined, **(b)** female, and **(c)** male models. Each column represents a model fold. Left eye averages are shown on upper rows (L), right eye averages on lower rows (R).

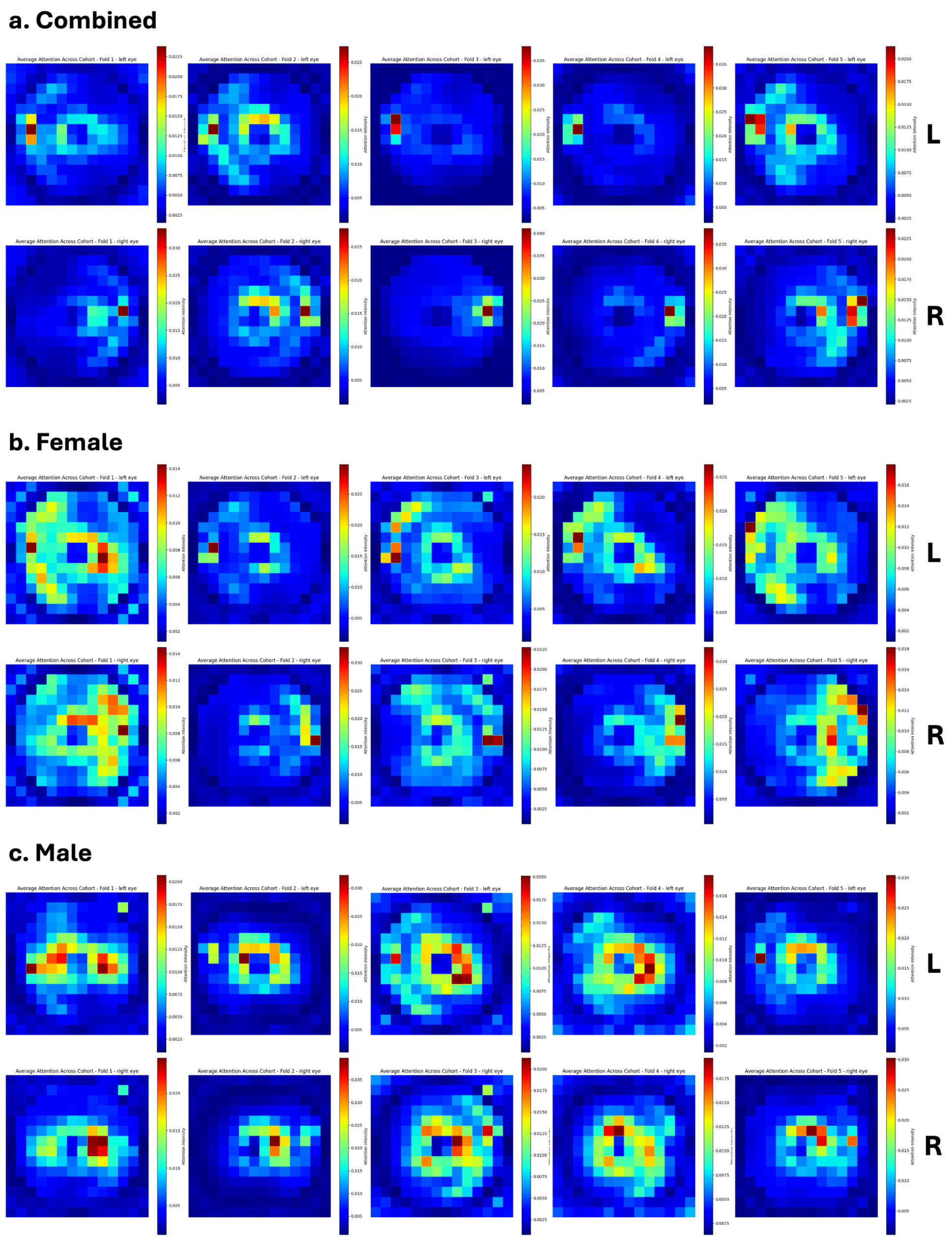


**Figure S6. Group-average patch-level attention maps in the 5 folds of the fine-tuned models.** Attention maps, averaged across heads and subjects are shown for **(a)** the combined, **(b)** female, and **(c)** male models for left (L, upper rows) and right (R, lower rows). Each column represents the last attention block of a model fold.

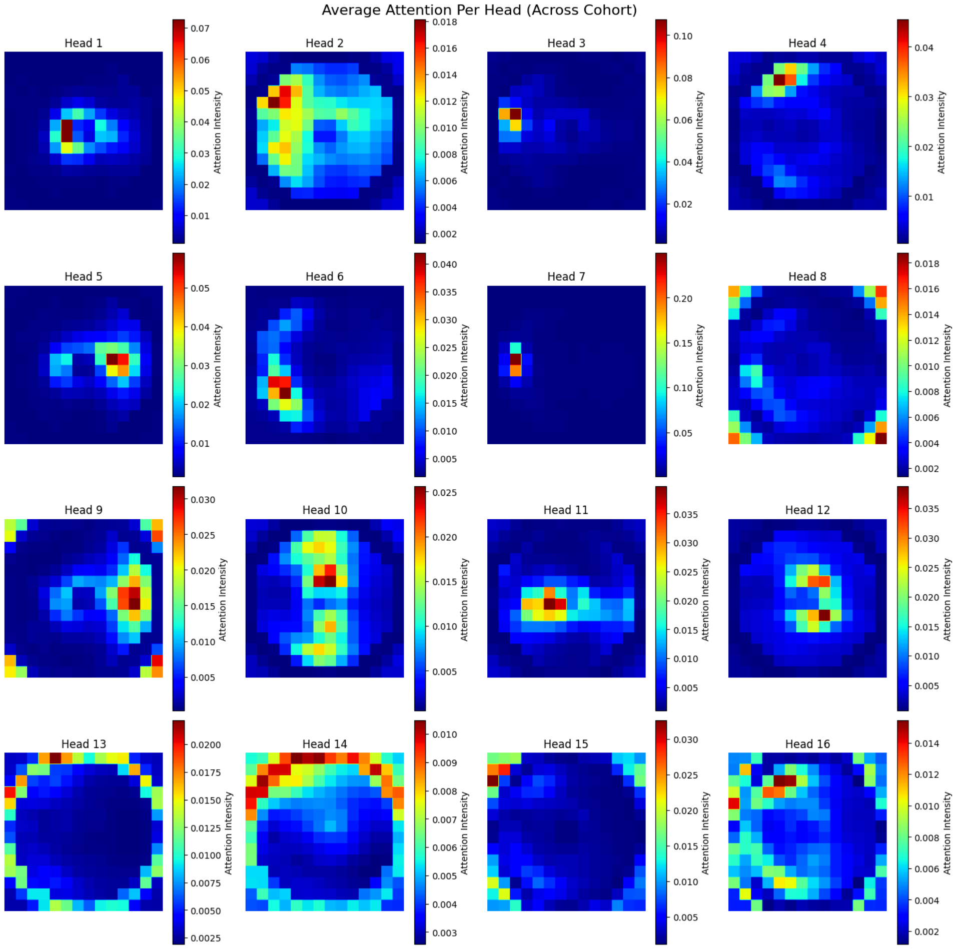


**Figure S7. Group-average left eye attention in separate heads of the combined model's fold 1, last attention block.**

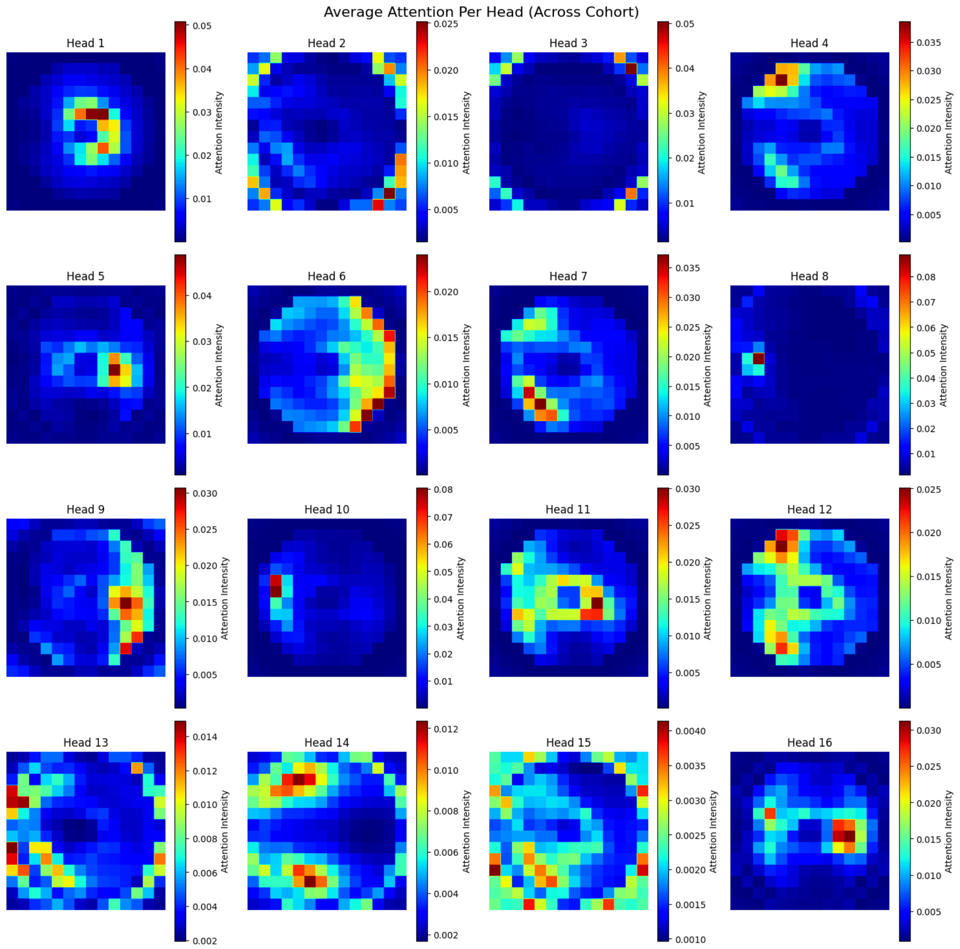


**Figure S8. Group-average left eye attention in separate heads of the female model's fold 1, last attention block.**

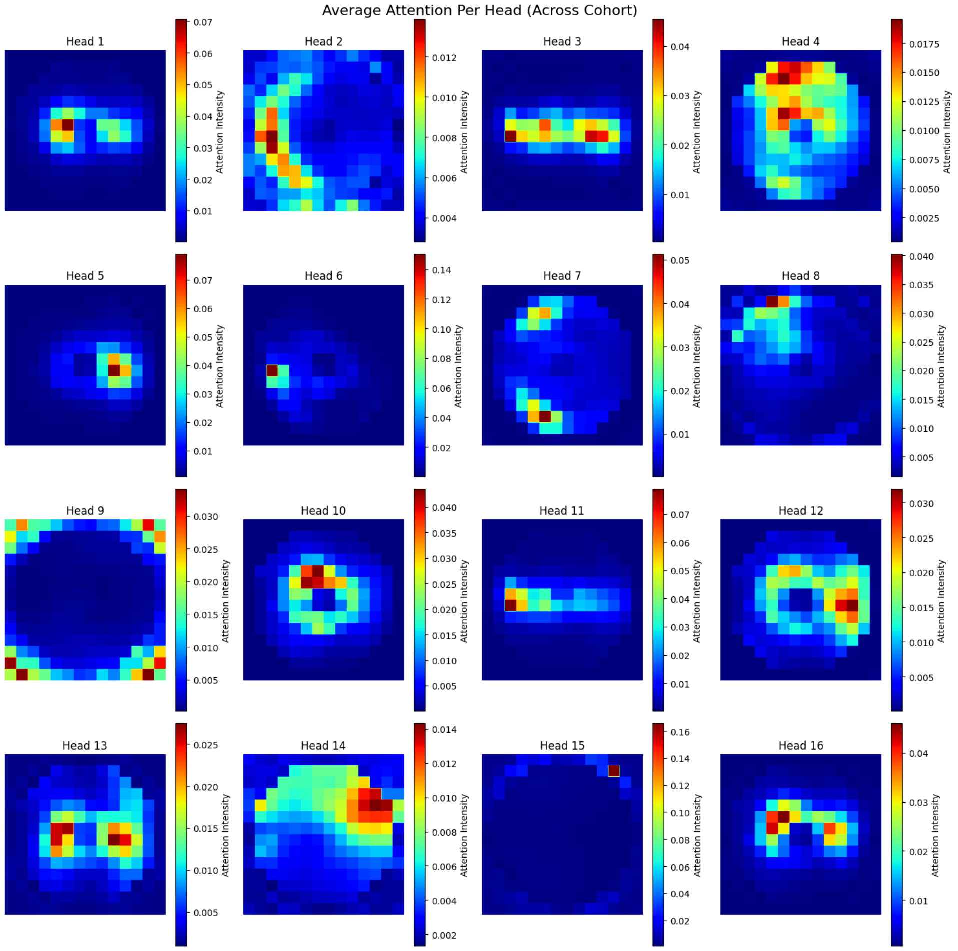


**Figure S9. Group-average left eye attention in separate heads of the male model's fold 1, last attention block.**

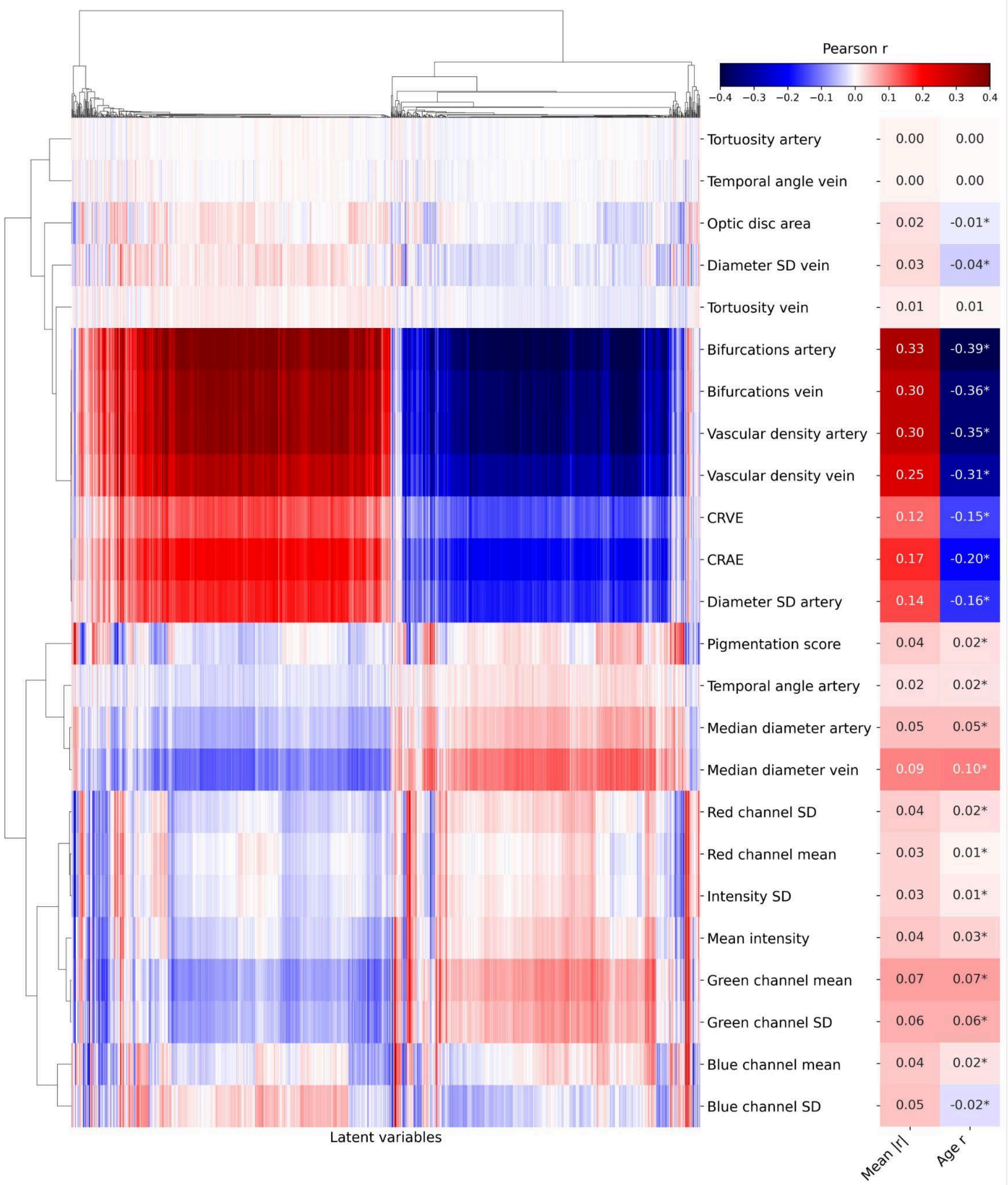


**Figure S10**. **Fold 2 correlation map between the combined model's 1024 latent variables and interpretable image features.** For every image feature, the mean absolute correlation with latent variables and the correlation with chronological age are shown on the right side of the figure. CRAE: central retinal arterial equivalent, CRVE: central retinal venous equivalent, SD: standard deviation, *: $p$<0.05.

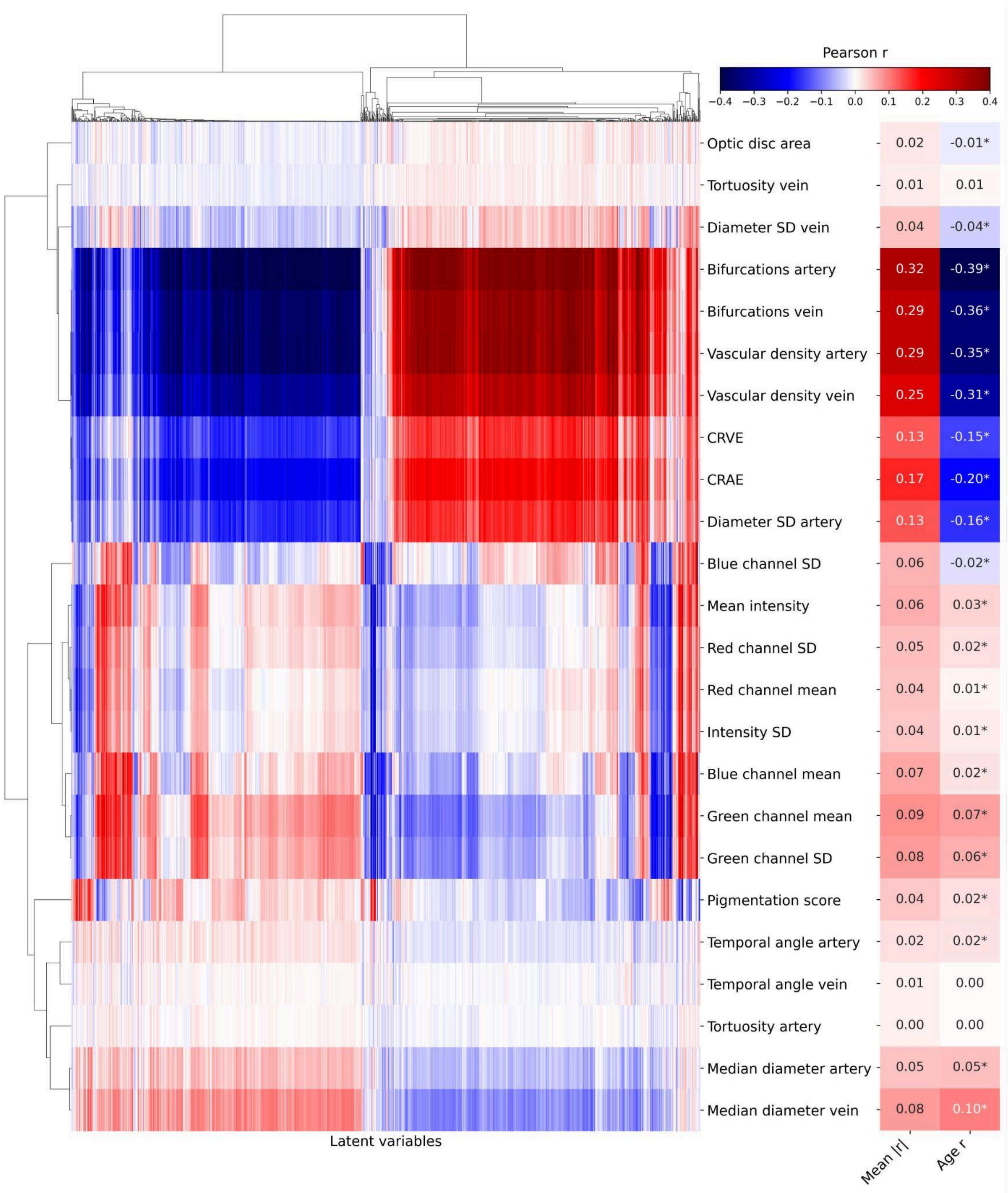


**Figure S11**. **Fold 3 correlation map between the combined model's 1024 latent variables and interpretable image features.** For every image feature, the mean absolute correlation with latent variables and the correlation with chronological age are shown on the right side of the figure. CRAE: central retinal arterial equivalent, CRVE: central retinal venous equivalent, SD: standard deviation, *: *p*<0.05.

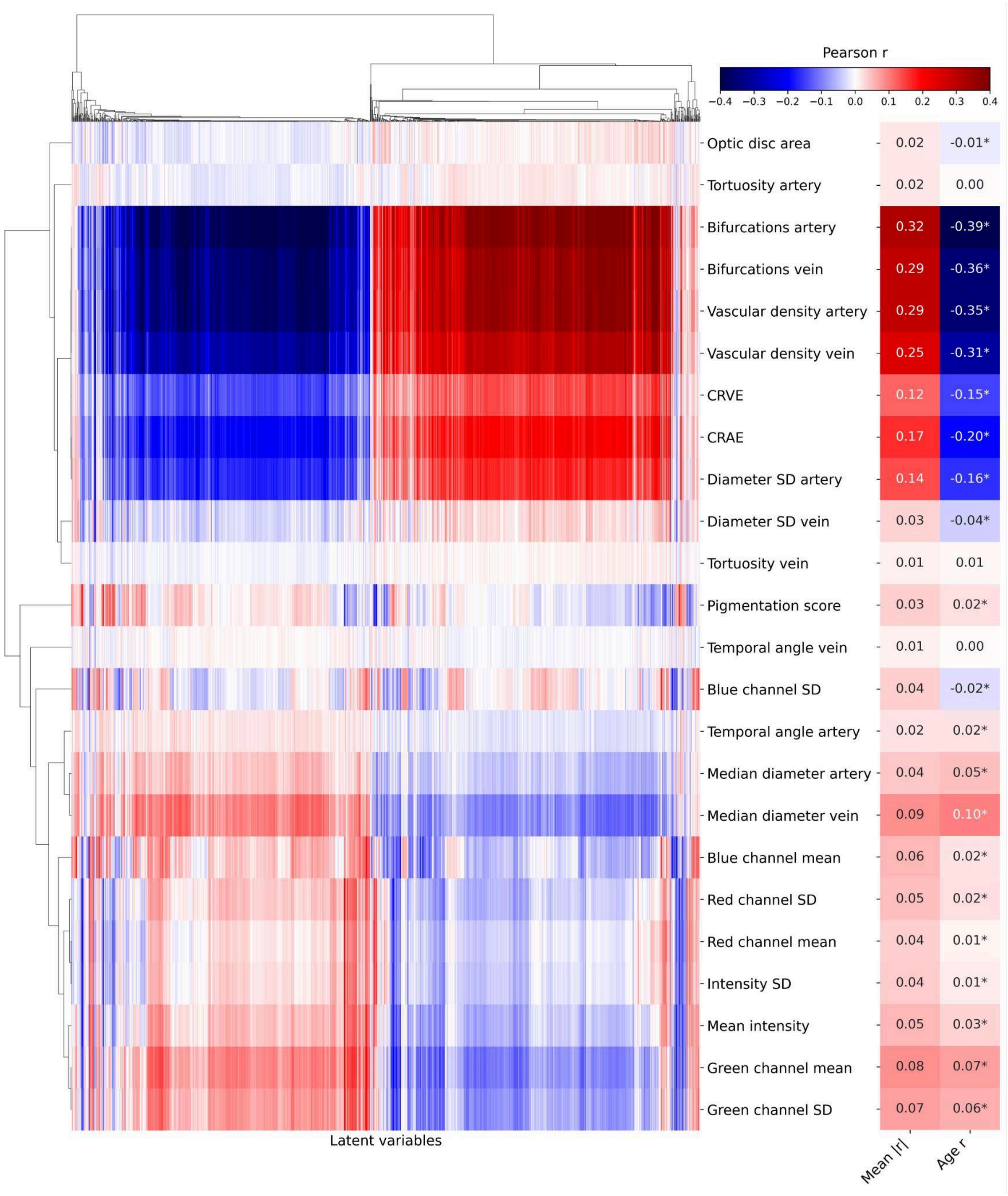


**Figure S12**. **Fold 4 correlation map between the combined model's 1024 latent variables and interpretable image features.** For every image feature, the mean absolute correlation with latent variables and the correlation with chronological age are shown on the right side of the figure. CRAE: central retinal arterial equivalent, CRVE: central retinal venous equivalent, SD: standard deviation, *: $p$<0.05.

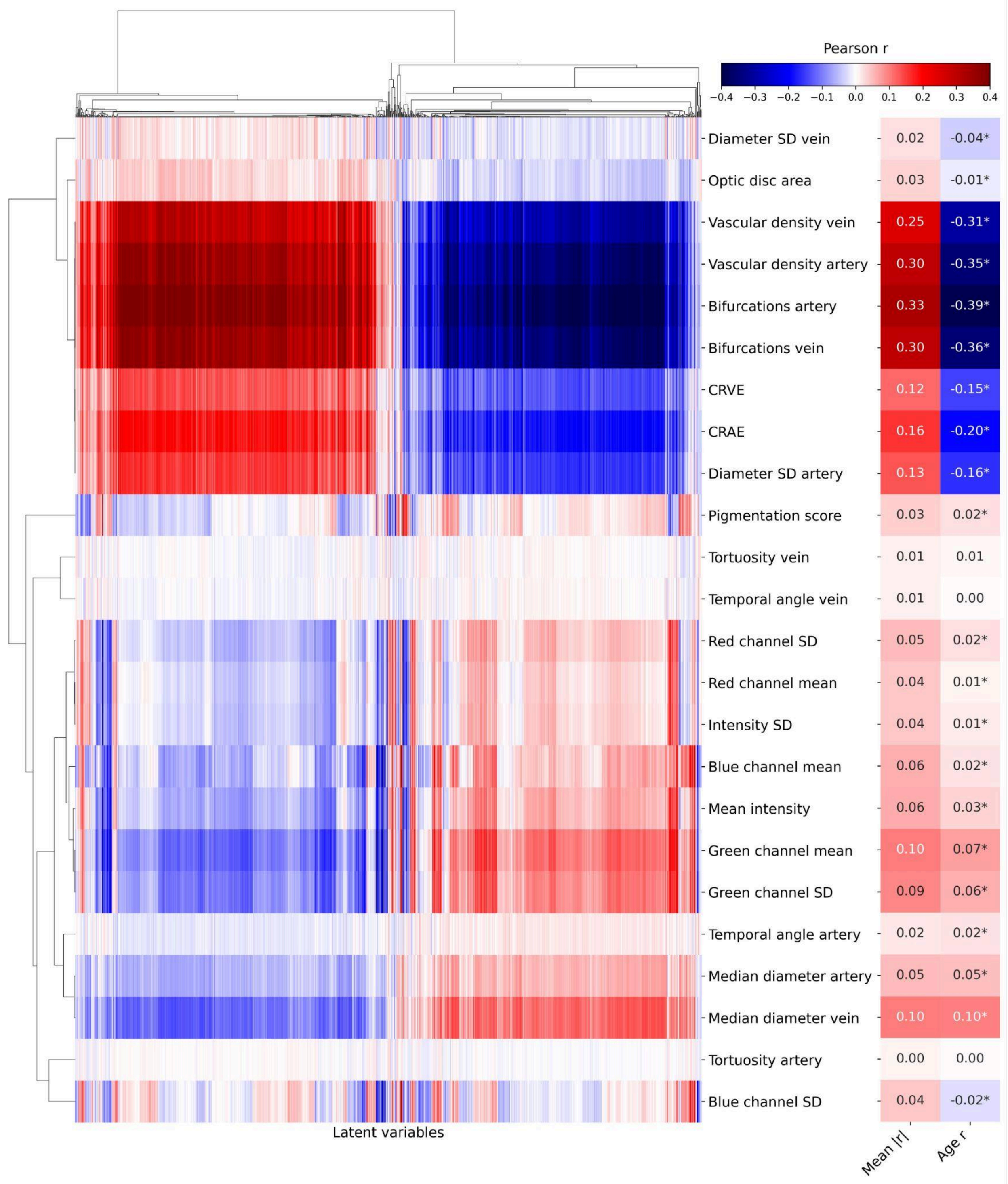


**Figure S13**. **Fold 5 correlation map between the combined model's 1024 latent variables and interpretable image features.** For every image feature, the mean absolute correlation with latent variables and the correlation with chronological age are shown on the right side of the figure. CRAE: central retinal arterial equivalent, CRVE: central retinal venous equivalent, SD: standard deviation, *: $p$<0.05.

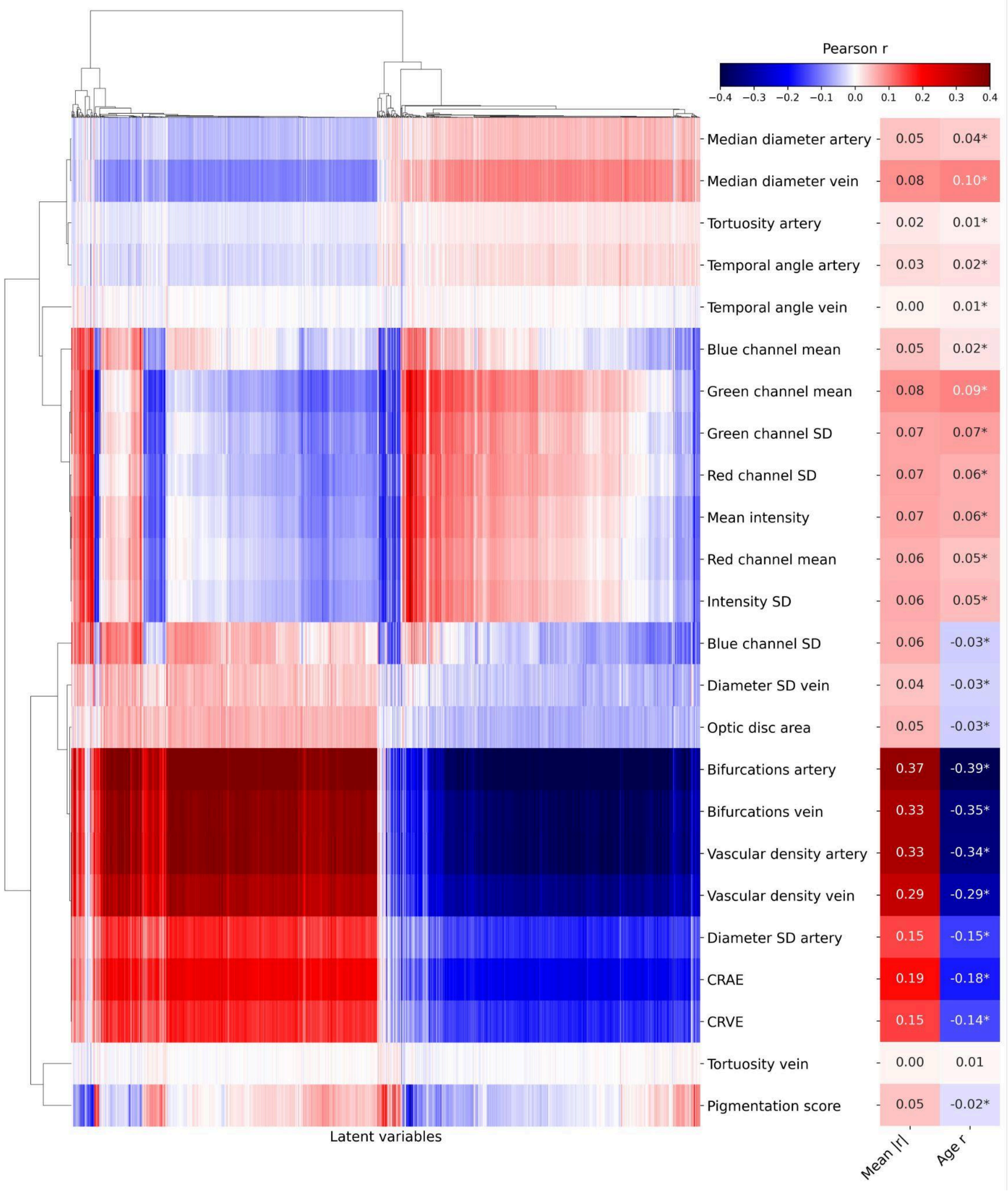


**Figure S14**. **Fold 1 correlation map between the female model's 1024 latent variables and interpretable image features.** For every image feature, the mean absolute correlation with latent variables and the correlation with chronological age are shown on the right side of the figure. CRAE: central retinal arterial equivalent, CRVE: central retinal venous equivalent, SD: standard deviation, *: $p$<0.05.

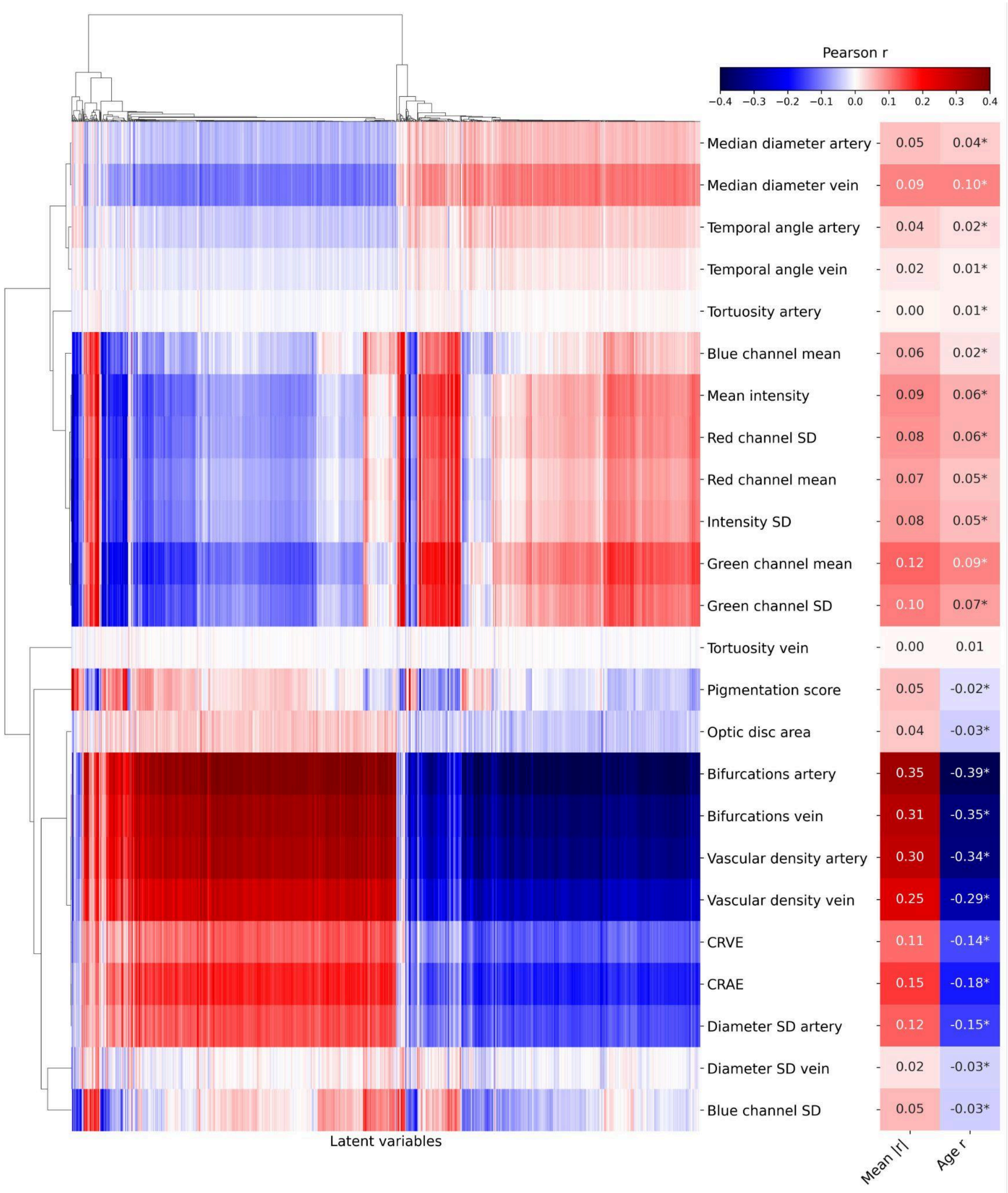


**Figure S15**. **Fold 2 correlation map between the female model's 1024 latent variables and interpretable image features.** For every image feature, the mean absolute correlation with latent variables and the correlation with chronological age are shown on the right side of the figure. CRAE: central retinal arterial equivalent, CRVE: central retinal venous equivalent, SD: standard deviation, *: $p$<0.05.

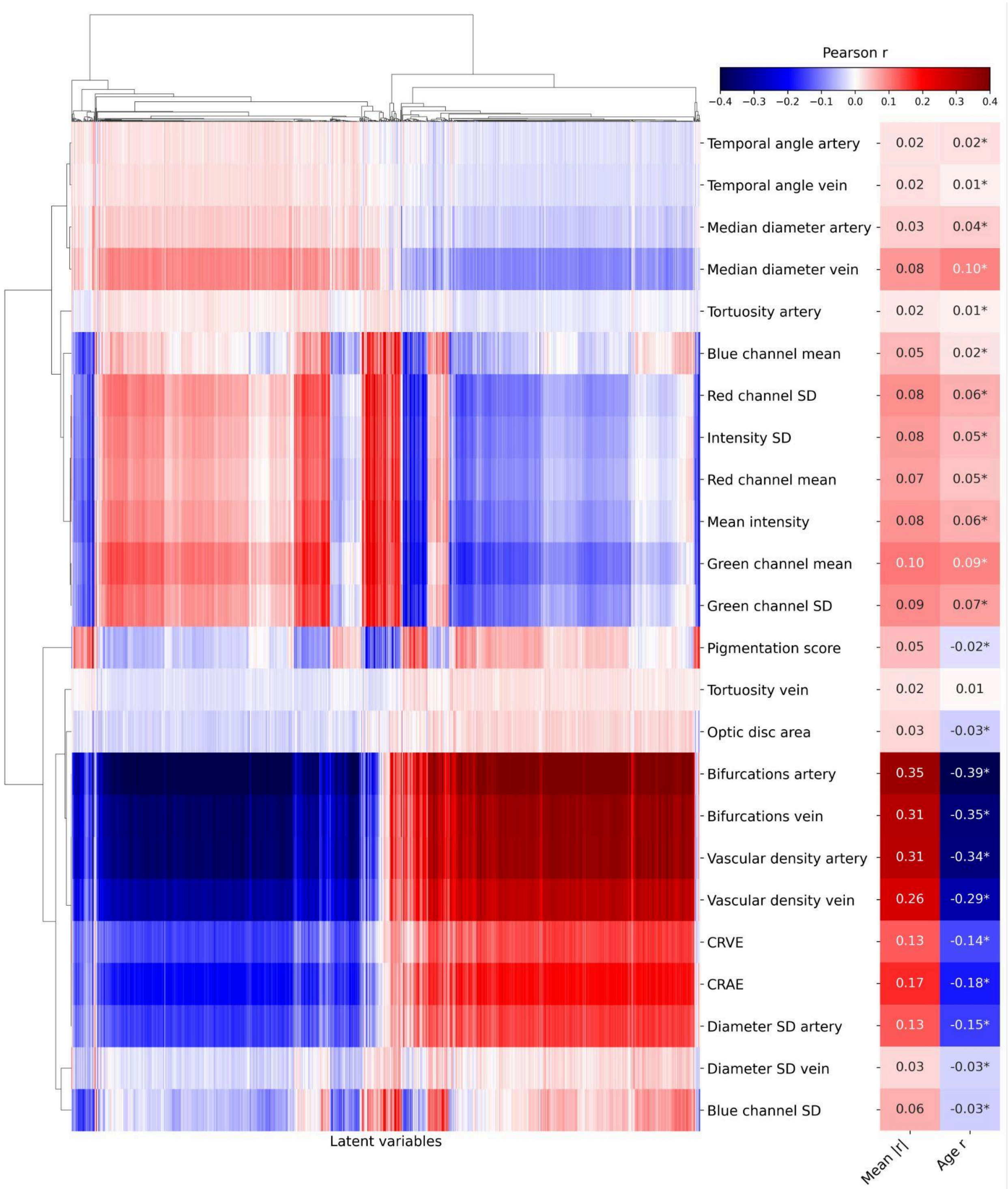


**Figure S16**. **Fold 3 correlation map between the female model's 1024 latent variables and interpretable image features.** For every image feature, the mean absolute correlation with latent variables and the correlation with chronological age are shown on the right side of the figure. CRAE: central retinal arterial equivalent, CRVE: central retinal venous equivalent, SD: standard deviation, *: *p*<0.05.

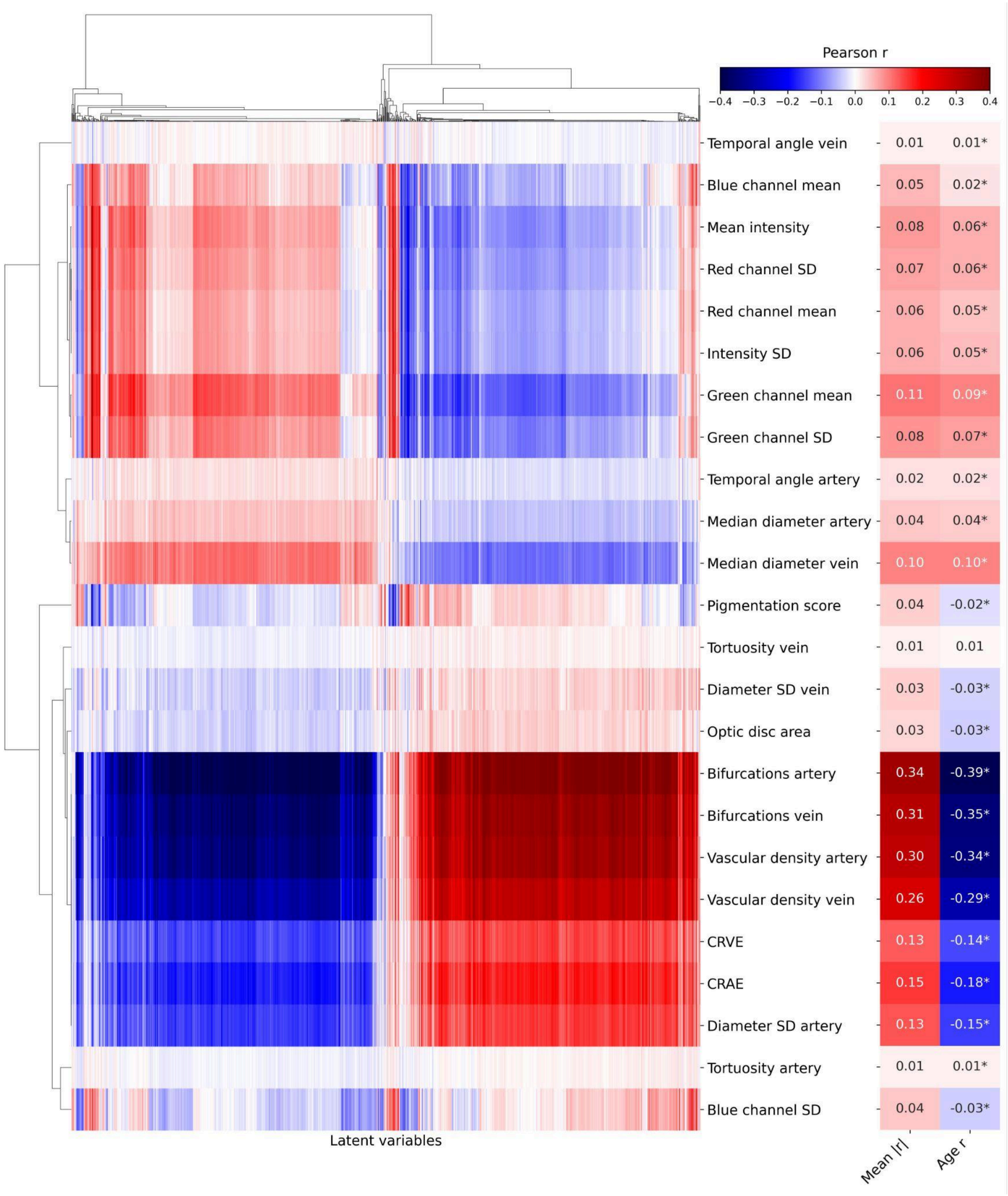


**Figure S17. Fold 4 correlation map between the female model's 1024 latent variables and interpretable image features.** For every image feature, the mean absolute correlation with latent variables and the correlation with chronological age are shown on the right side of the figure. CRAE: central retinal arterial equivalent, CRVE: central retinal venous equivalent, SD: standard deviation, *: $p<0.05$.

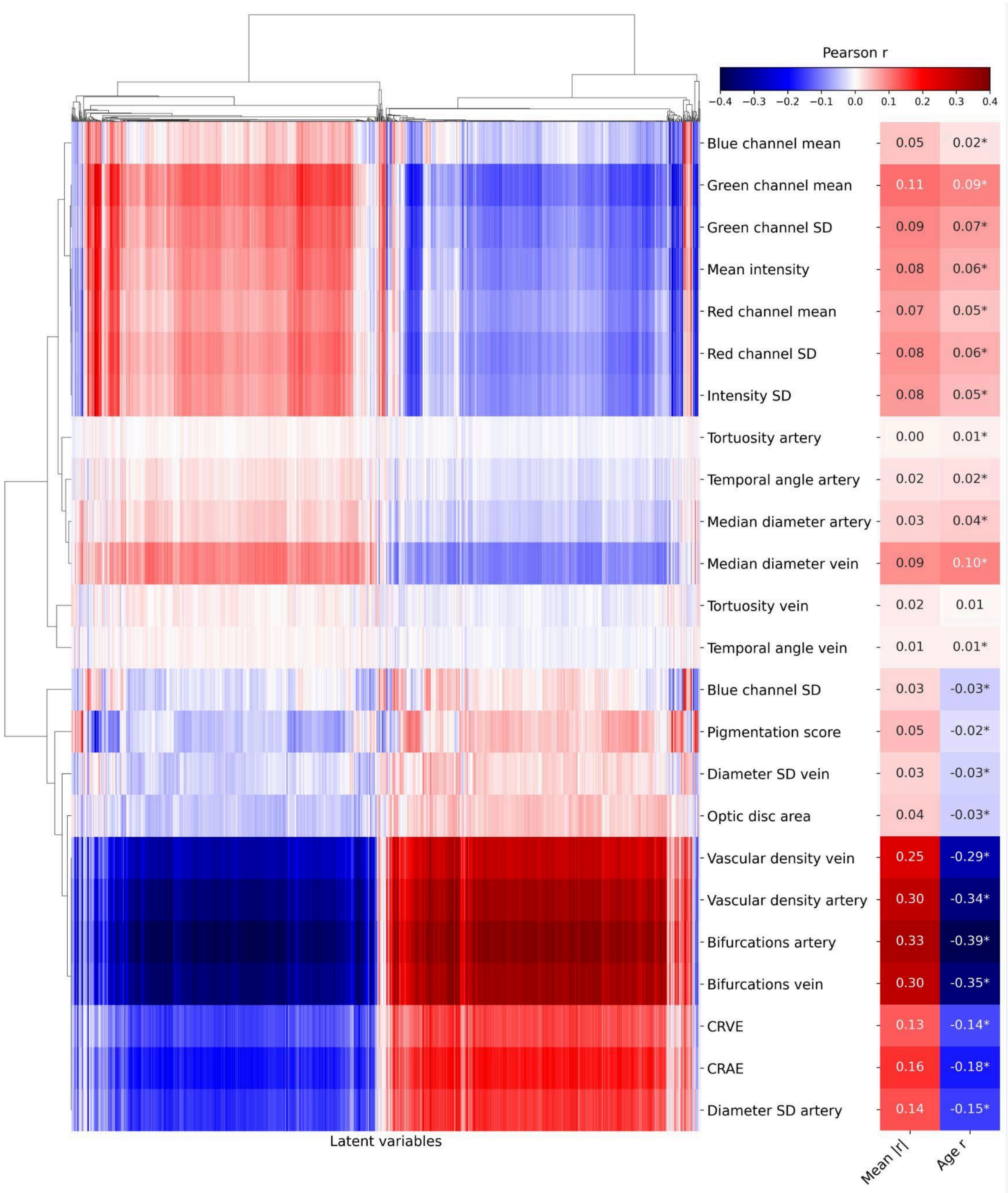


**Figure S18**. **Fold 5 correlation map between the female model's 1024 latent variables and interpretable image features.** For every image feature, the mean absolute correlation with latent variables and the correlation with chronological age are shown on the right side of the figure. CRAE: central retinal arterial equivalent, CRVE: central retinal venous equivalent, SD: standard deviation, *: $p$<0.05.

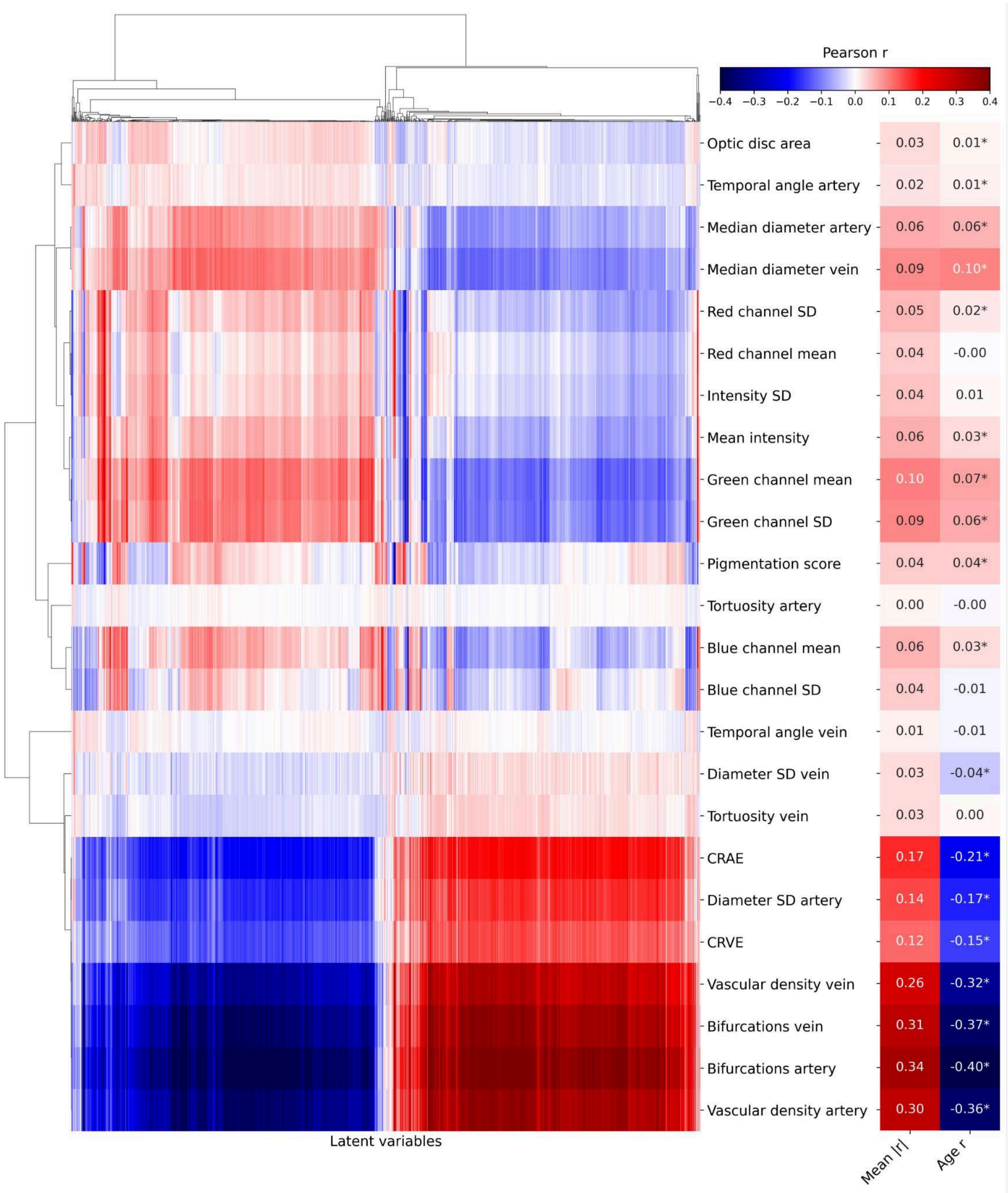


**Figure S19**. **Fold 1 correlation map between the male model's 1024 latent variables and interpretable image features.** For every image feature, the mean absolute correlation with latent variables and the correlation with chronological age are shown on the right side of the figure. CRAE: central retinal arterial equivalent, CRVE: central retinal venous equivalent, SD: standard deviation, *: $p$<0.05.

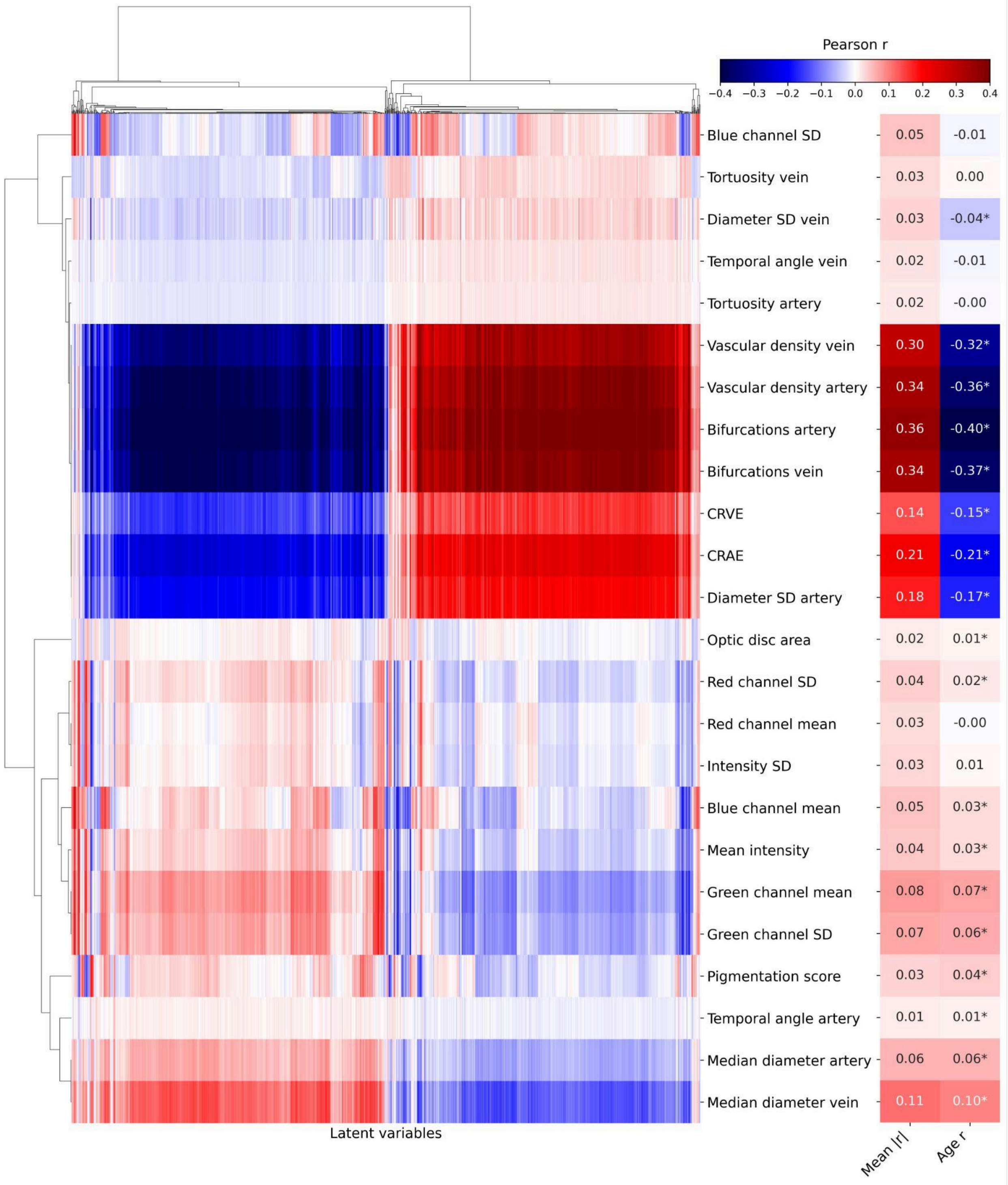


**Figure S20**. **Fold 2 correlation map between the male model's 1024 latent variables and interpretable image features.** For every image feature, the mean absolute correlation with latent variables and the correlation with chronological age are shown on the right side of the figure. CRAE: central retinal arterial equivalent, CRVE: central retinal venous equivalent, SD: standard deviation, *: $p$<0.05.

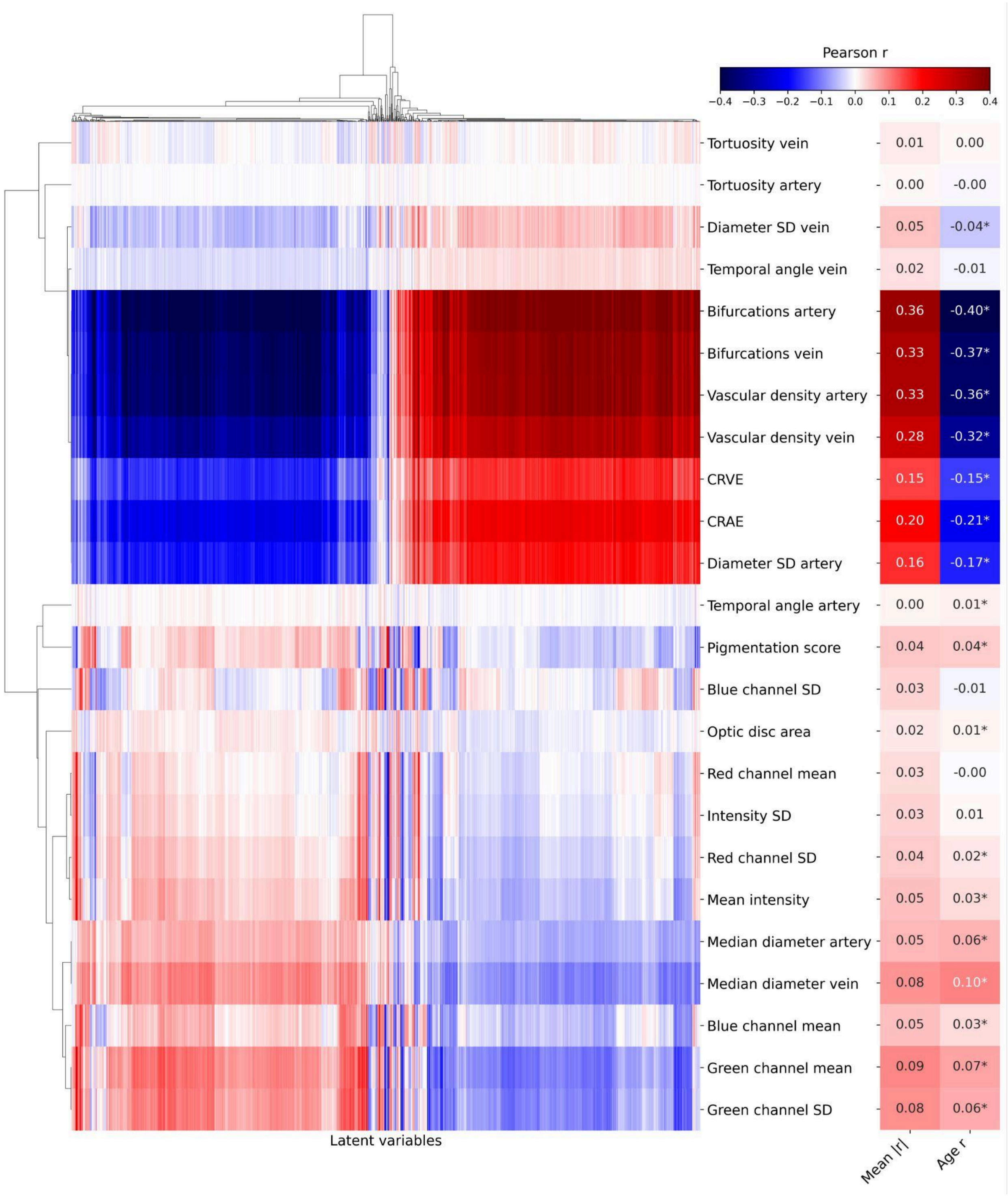


**Figure S21**. **Fold 3 correlation map between the male model's 1024 latent variables and interpretable image features.** For every image feature, the mean absolute correlation with latent variables and the correlation with chronological age are shown on the right side of the figure. CRAE: central retinal arterial equivalent, CRVE: central retinal venous equivalent, SD: standard deviation, *: $p$<0.05.

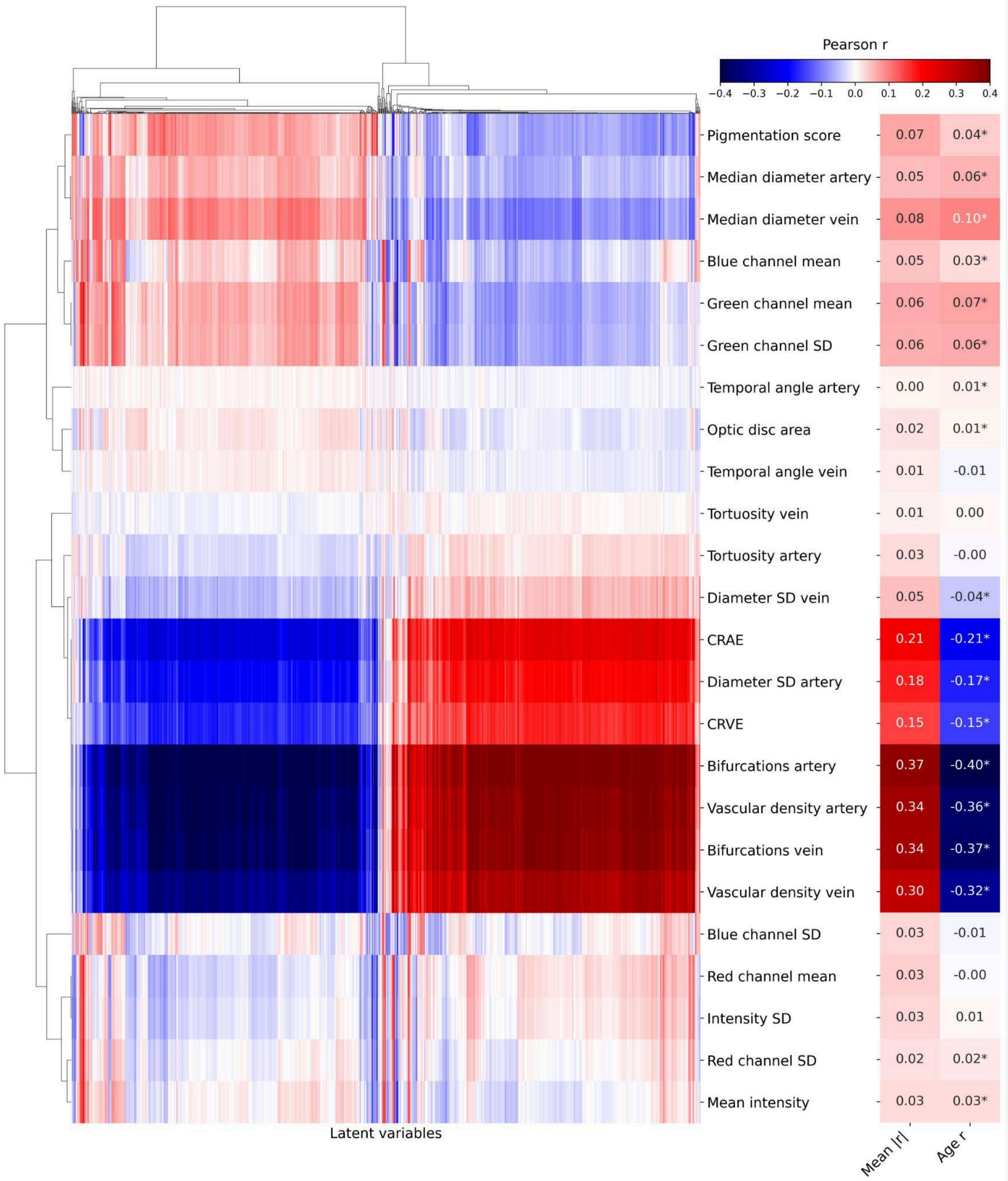


**Figure S22**. **Fold 4 correlation map between the male model's 1024 latent variables and interpretable image features.** For every image feature, the mean absolute correlation with latent variables and the correlation with chronological age are shown on the right side of the figure. CRAE: central retinal arterial equivalent, CRVE: central retinal venous equivalent, SD: standard deviation, *: $p$<0.05.

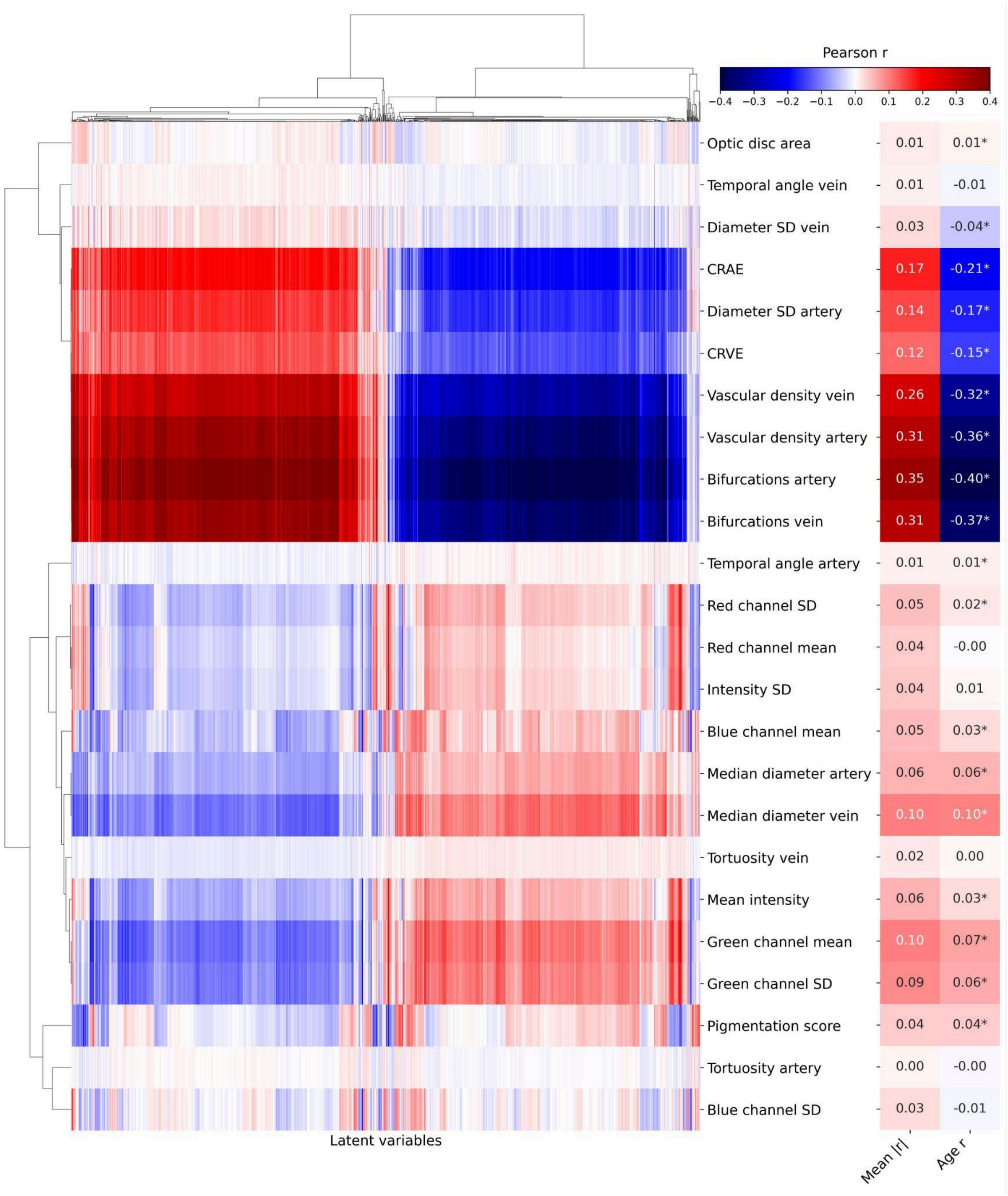


**Figure S23**. **Fold 5 correlation map between the male model's 1024 latent variables and interpretable image features.** For every image feature, the mean absolute correlation with latent variables and the correlation with chronological age are shown on the right side of the figure. CRAE: central retinal arterial equivalent, CRVE: central retinal venous equivalent, SD: standard deviation, *: $p<0.05$.
